# Mapping Cerebellar Morphology in 15q11.2 CNV Carriers Using Normative Modeling

**DOI:** 10.1101/2025.05.28.25328353

**Authors:** Milin Kim, Esten H. Leonardsen, Saige Rutherford, Geir Selbæk, Karin Persson, Nils Eiel Steen, Olav B. Smeland, Geneviève Richard, Dag Alnæs, Rune Boen, Ida E. Sønderby, Dennis van der Meer, Ole A. Andreassen, Lars T. Westlye, Thomas Wolfers, Torgeir Moberget

**Affiliations:** Centre for Precision Psychiatry, Division of Mental Health and Addiction, University of Oslo and Oslo University Hospital, Oslo, Norway; Department of Psychology, University of Oslo, Oslo, Norway; KG Jebsen Centre for Neurodevelopmental Disorders, University of Oslo, Oslo, Norway; Department of Behavioral Science, School of Health Sciences, Oslo Metropolitan University - OsloMet, Oslo, Norway; Department of Cognitive Neuroscience, Radboud University Medical Centre, Nijmegen, Netherlands; Donders Institute, Radboud University, Nijmegen, Netherlands; Department of Psychiatry, University of Michigan, Ann Arbor, MI, United States; Department of Psychiatry and Psychotherapy, University of Tübingen, Tübingen, Germany; German Center for Mental Health (DZPG), Tübingen, Germany; Department of Geriatric Medicine, Oslo University Hospital, Oslo, Norway; The Norwegian National Centre for Ageing and Health, Vestfold Hospital Trust, Tønsberg, Norway; Section for Clinical Psychosis Research, Division of Mental Health and Addiction, Oslo University Hospital, Oslo, Norway; Division of Mental Health and Substance Abuse, Diakonhjemmet Hospital, Oslo, Norway; Institute for clinical medicine, University of Oslo, Oslo, Norway; Department of Psychology, Pedagogy and Law, School of Health Sciences, Kristiania University College, Oslo, Norway; Department of Medical Genetics, Oslo University Hospital, Oslo, Norway; School of Mental Health and Neuroscience, Faculty of Health, Medicine and Life Sciences, Maastricht University, Maastricht 6200 MD, the Netherlands; Semel Institute for Neuroscience and Human Behavior, University of California Los Angeles, Los Angeles, CA, USA

**Keywords:** Cerebellum, Normative modelling, Magnetic Resonance Imaging, Copy Number Variations, Heterogeneity Mapping, 15q11.2

## Abstract

Copy number variations (CNVs) at the 15q11.2 locus of the human genome have been associated with altered brain structure and increased risk for neurodevelopmental and neuropsychiatric disorders. The cerebellum is increasingly seen as a crucial brain region for neurodevelopmental conditions, yet the effects of 15q11.2 CNVs on cerebellar morphology remain largely unclear. Importantly, 15q11.2 CNVs shows reduced or incomplete penetrance (meaning that not all CNV carriers are affected) and variable expressivity (meaning that symptoms may differ between individuals with the same genetic alteration). Thus, there is a need to not only assess group differences, but also to quantify anatomical variability at the individual level. Here, we address these issues using normative models of brain anatomy trained on large datasets (n > 52k, age range: 3-85) to assess both group and individual-level deviations in cerebellar anatomy in carriers of 15q11.2 deletions (n = 120, mean [SD] age= 64.95 [7.58]) and duplications (n = 149, mean [SD] age=64.31 [7.21]), compared to non-carriers (n = 19,028, mean [SD] age=64.31 [7.58]). Group-level case-control analyses revealed significantly smaller total and regional cerebellar volumes in both deletion and duplication carriers, though with small effect sizes. Individual-level deviation analyses, capturing pronounced alterations in specific individuals, revealed a heterogeneous pattern among carriers. Overall, our findings suggest that CNVs at the 15q11.2 locus exert modest and highly individualized effects on cerebellar morphology.

## Introduction

Pathogenic copy number variations (CNVs) refer to deletions or duplications of longer stretches of genetic material that have been linked to increased risk of developing a disorder or disease (Fig. 1A) (Kirov, 2015; Takumi & Tamada, 2018). Since such CNVs span a limited number of genes, they may provide a unique window onto the mechanisms linking genetic risk to pathophysiology. For neurodevelopmental disorders such mechanisms are likely to involve brain development and morphology (Mollon et al., 2023; Sønderby et al., 2022). For instance, the very rare 22q11.2 deletion (affecting 1 in 2000 live births (Fung et al., 2015)) is linked not only to a 25-fold increased risk for schizophrenia (SZ) (Leslie et al., 2024; Owen et al., 2023) and a 10-fold increased risk for autism spectrum disorders (ASD) (Clements et al., 2017), but also to altered brain morphology as measured by Magnetic Resonance Imaging (MRI; Schmitt et al., 2023). More common pathogenic CNVs linked to neurodevelopmental disorders, such as CNVs at the 15q11.2 locus, which confer lower disease risk, have also been associated with altered cognitive function, brain morphology as well as developmental and motor delay (Cafferkey et al., 2014; Doornbos et al., 2009; Stefansson et al., 2014; Ulfarsson et al., 2017; Writing Committee for the ENIGMA-CNV Working Group et al., 2019) while also increasing the susceptibility of developing neurodevelopmental and/or psychiatric disorders, such as SZ (Cafferkey et al., 2014; Cook Jr & Scherer, 2008; Stefansson et al., 2008; Vaez et al., 2023), attention-deficit/hyperactivity disorder (ADHD) (Gudmundsson et al., 2019), and ASD (Cox & Butler, 2015). Situated in an unstable genetic region that is susceptible to variations, often occurring at specific chromosomal breakpoints (BPs) (Busch et al., 2023), the prevalence of 15q11.2 locus is 0.44% for deletion and 0.49% in duplication (Calle Sánchez et al., 2022).

**Fig. 1.**
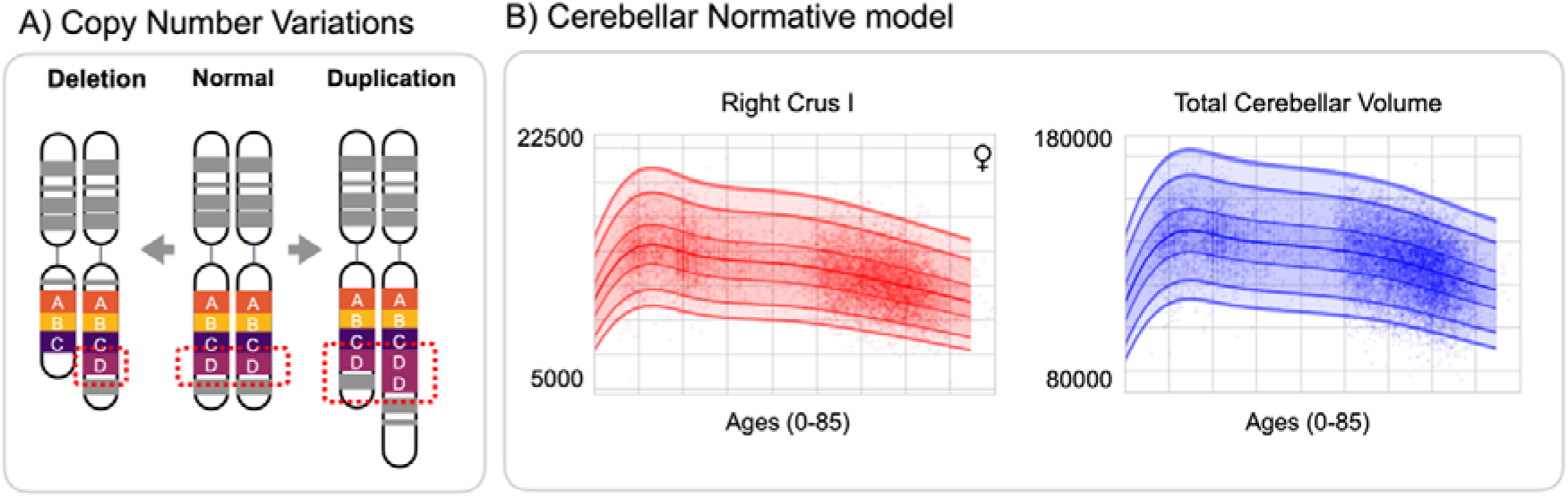
The cerebellar normative model without CNV carriers. (A) CNV refers to variations in the number of genome copies of a certain locus, which can be either deleted (only one copy) or duplicated (three or more copies). (B) Displayed is the cerebellar normative model, excluding CNV carriers from the testing set. Two of the 28 regions of the cerebellum, including the total cerebellar volume, are shown to illustrate the lobular growth chart. The *x*-axis represents the age range, while the *y*-axis indicates the predicted cerebellar volume for each sex. Lines denote the centiles, while dots represent individual participants.

Numerous studies have found that individuals carrying these CNVs exhibit incomplete or reduced penetrance and variable expressivity of the associated clinical phenotype (Kingdom & Wright, 2022), meaning that not all CNV carriers are affected and that symptoms may differ between individuals with the same genetic alteration. For neuropsychiatric outcomes, the penetrance of 15q11.2 deletions and duplications have been estimated to be between 7.3% and 6.28% (Maya et al., 2020). Moreover, “gene dosage” effect can potentially result in corresponding dosage effects on the measured phenotype, where carriers of deletions (1 copy) and duplications (3 copies) show opposing effects relative to non-carriers (2 copies) (Feuk et al., 2006; Kendall et al., 2017). Previous MRI studies of the 15q11.2 CNV have reported such gene dosage effects for a limited number of brain phenotypes, namely cerebro-cortical thickness (Writing Committee for the ENIGMA-CNV Working Group et al., 2019), and white matter microstructure (Silva et al., 2021), while the cerebellum has received significantly less attention. This omission is surprising, since cerebellum has also been linked to neurodevelopmental/psychiatric disorders such as ASD and SZ (Hughes et al., 2023; Kim et al., 2024), Indeed, for the rare 22q11.2 deletions, which strongly increase the risk for ASD and SZ (Fiksinski et al., 2023), a meta-analysis found that smaller total cerebellar volume was the most pronounced brain structural alteration associated with 22q11.2 deletions, with a Hedge’s g effect size of −1.25 (Rogdaki et al., 2020). A more recent study also reported reduced volumes of cerebellar lobules VII and VIII in 22q11.2 deletions (Schmitt et al., 2023).

Here, we comprehensively map cerebellar and cerebral morphology in 15q11.2 deletions and duplications through normative modeling (Kim et al., 2024; Marquand et al., 2016, 2019; Rutherford, Fraza, et al., 2022; Rutherford et al., 2023; Wolfers et al., 2018, 2021; Zabihi et al., 2019). Using these models, we evaluated deviations from the population reference in carriers of deletions (n = 120) and duplications (n=149) at the 15q11.2 locus and compared these to the deviations in non-carriers (n =19,028). Our research advances the existing literature by i) estimating the effect of this CNV on cerebellar morphology at the group level, and ii) exploring the utility of normative models in charting the distribution of extreme deviations within each group and individual. Given the known reduced penetrance and variable expressivity of CNVs (Maya et al., 2020; Rosenfeld et al., 2013) at the 15q11.2 locus, we anticipated that while the majority of carriers would fall within the normal range for cerebellar morphology, a larger proportion of carriers than non-carriers would still exhibit significant deviations from the population reference.

## Results

To quantify individual variation in cerebellar anatomy, we employed normative modeling to calculate deviations (z-scores) from the population reference. The cerebellar normative models were developed using cerebellar features as illustrated in Figure 1B and Supplementary Figure 1. Note that these z-scores are automatically adjusted for effects of age, sex- and scanner type. In addition to directly comparing mean z-scores across groups, we also identified individuals with pronounced deviations from the population reference by defining extreme deviations as z-scores with (|*z*|>1.96).

Effects of CNV status on mean z-scores for total cerebellar volume and 28 cerebellar regional volumes (see Supplementary Table 2) were tested for using one-way ANOVAs (three levels: deletions, non-carriers, duplications). Figure 2A shows distribution of z-scores for total cerebellar volume across groups, while Figure 2B shows the estimated carrier group means (deletion, duplication and non-carriers) and their confidence intervals. The ANOVA confirmed a significant effect of carrier group (*F*=7.34, *p*=.0006, η²=0.0007) (Supplementary Table 2), while follow-up pair-wise comparisons showed that total cerebellar volume was lower in both deletion-carriers (T= −2.73, *p*=0.007, Cohen’s d= −0.27) and duplication-carriers (T= −2.23, *p*=0.027, Cohen’s d= −0.21) relative to non-carriers (see also Supplementary Table 3 and Figure 2B).

**Fig. 2.**
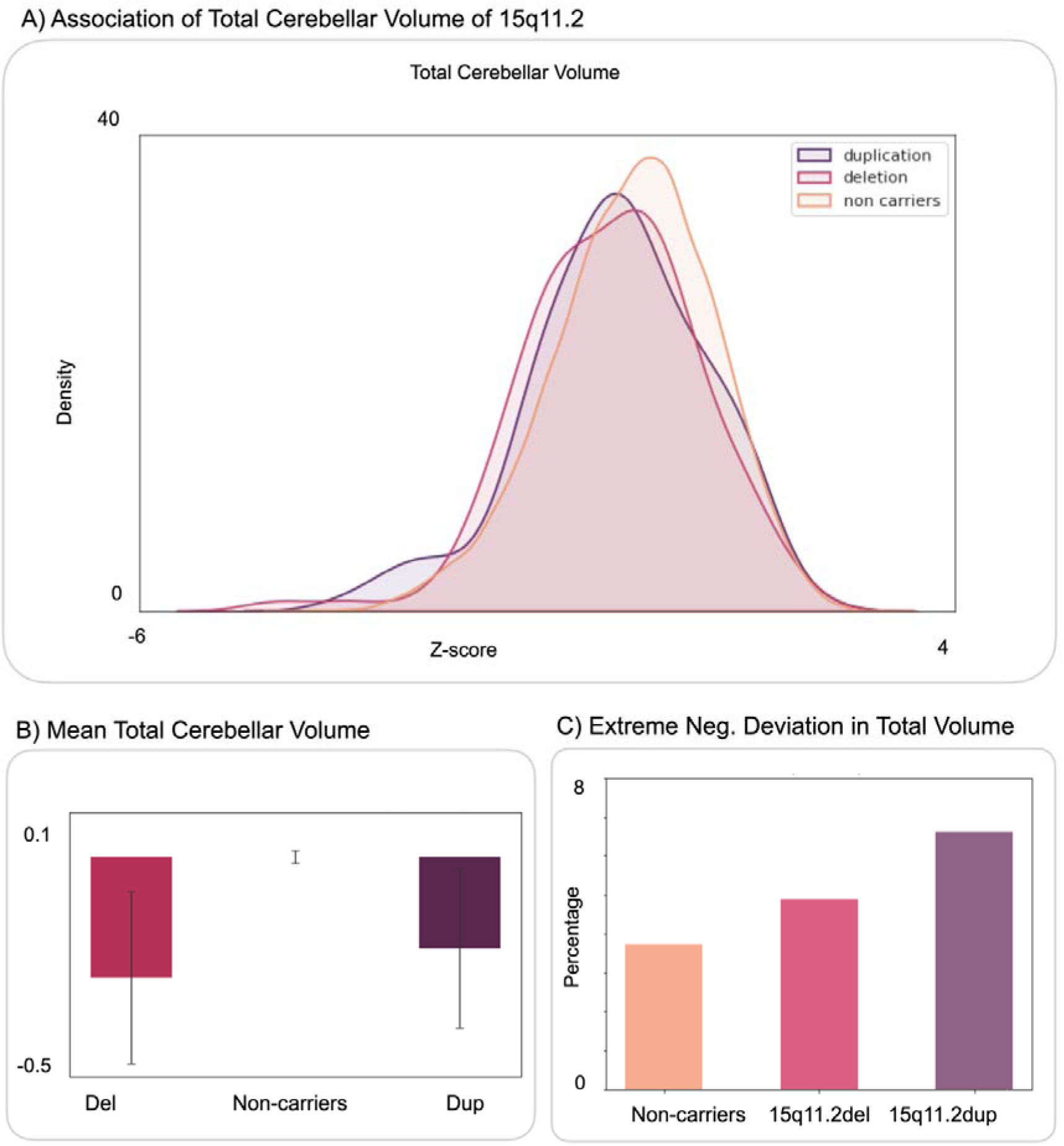
The 15q11.2 show significant reduction in total cerebellar volume compared to non-carriers. (A) This figure shows distributions of total cerebellar volume across carrier groups. (B) Individuals with 15q11.2 deletions (Cohen’s d= −0.27) and duplications (Cohen’s d= −0.21) show significant reductions in total cerebellar volume compared to non-carriers. (C) Both 15q11.2 duplications and deletions exhibit a higher proportion (in percentage) of extreme negative deviations in total volume

Figure 2C displays the percentage of extreme negative and positive deviations in total cerebellar volume (i.e., |*z*|L>1.96). Both deletion (5.0%) and duplication (6.71%) carriers tended to show a slightly higher proportion of individuals with extreme negative deviations than non-carriers (3.72%). A chi-square test for independence was conducted to examine the relationship between the proportion of extreme deviations in the total cerebellar volume and the carrier groups. The results showed no significant association between these two variables (χ²(4) = 4.91, *p*=0.30) (Supplementary Table 5). Note also that all three groups showed larger percentages of extreme negative than positive deviations in total cerebellar volume, possibly due to relatively advanced age of the current sample.

Regional ANOVAs of the 28 cerebellar lobules revealed significant effects of carrier group in several regions (Supplementary Table 2). Follow-up paired t-tests revealed significant volume reductions in right Crus I (T= −3.35, uncorrected *p* =0.001, Cohen’s d = −0.32) and Vermis VII (T=-3.23, uncorrected *p*=0.002, Cohen’s d= −0.30) for the deletion carriers. For duplication carriers, reductions were shown in right IV (T= −3.80, corrected, uncorrected *p*=0.000, Cohen’s d = −0.30) and left VI (T= −3.12, uncorrected *p* =0.002; Cohen’s d= −0.257) relative to the non-carriers (see lower row in Figure 3A; for completeness we also show nominally significant (uncorrected) pair-wise comparisons in the upper row).

**Fig. 3.**
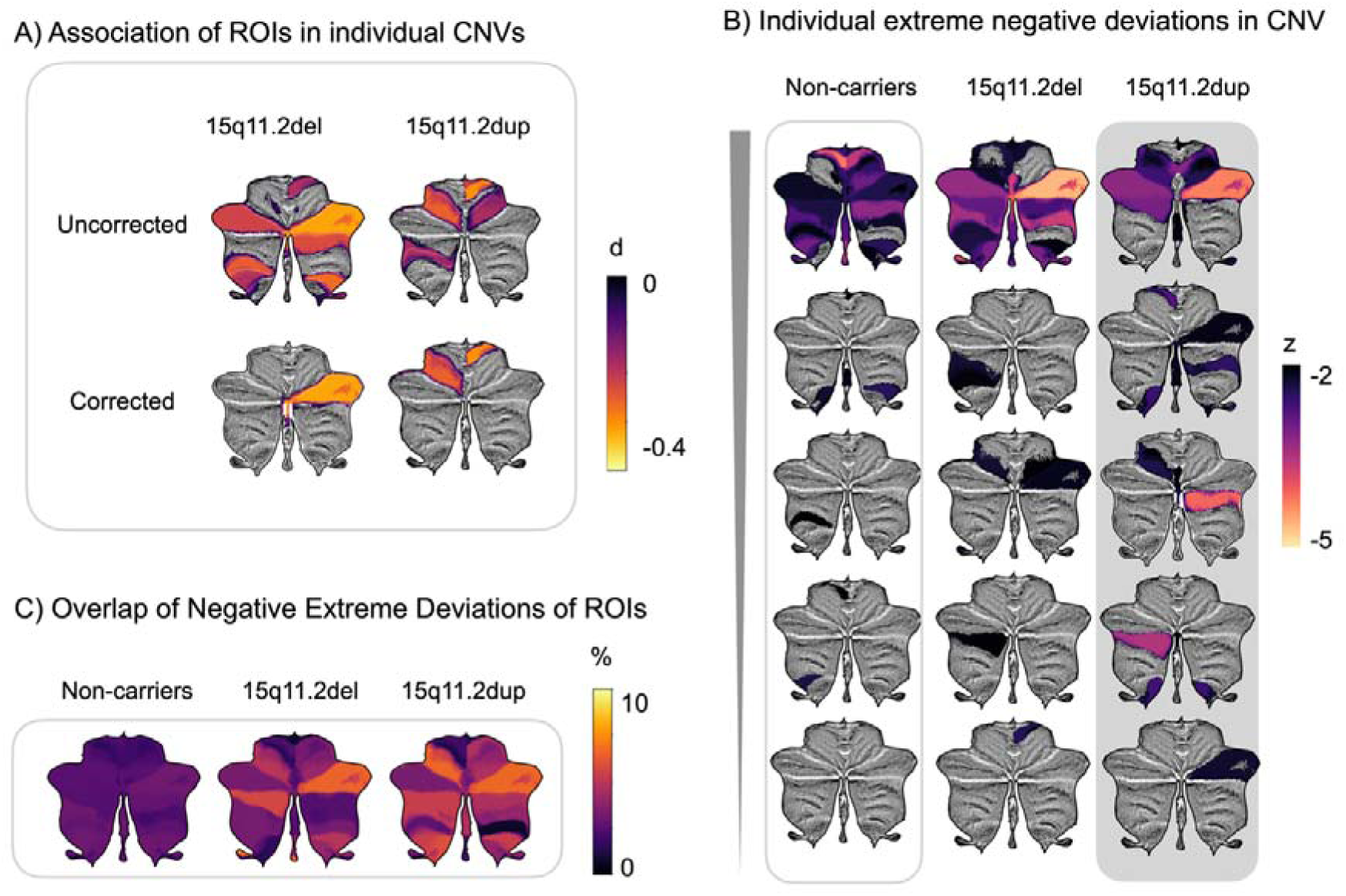
The 15q11.2 exhibits a moderate association with significant deviations in the cerebellar lobules. (A) Notably, the Right Crus I and Vermis VII are primarily associated with deletions, whereas the Right IV and Left VI are linked to duplications, showing no overlaps between the 15q11.2 carriers. (B) This illustrates the overlap of extreme negative deviations within a cohort, with the percentages calculated for each lobule. (C) Extreme negative deviations in non-carriers and individuals with CNVs exhibit unique patterns across different lobules.

Figure 3B gives examples of regional extreme negative deviations in five randomly selected individuals from each quintile of each group. The quintiles were established by arranging individuals in ascending order based on the number of extreme positive or negative deviations. Figure 3C shows the overlap of such extreme negative deviations in each group (i.e., the percentage of group members showing extreme negative deviations in each region; see Supplementary Table 4). The highest percentages of extreme negative deviations were found in the following regions: 15q11.2 deletion in Vermis X (8.33%), 15q11.2 duplication in the Right Crus I (6.71%) and Left VI (6.71%) and Right Crus I in non-carriers (3.41%). When comparing percentage of extreme negative and positive deviations in regional cerebellar volume using chi-square test, we found that there were significant associations in several regions (Supplementary Table 5). In the post-hoc test, we found significant group difference in extreme negative deviations between 15q11.2 deletion and non-carriers in Vermis X (Z-score =3.45, uncorrected p=.0006) and left VI for duplications (Z-score =3.11, uncorrected p=.0019) (Supplementary Table 6). Analyses using voxel-wise cerebellar z-scores did not yield any significant results after multiple comparisons correction (see Supplementary Figure 3A-C).

To directly compare the cerebellar results of the current study with previously reported cerebral effects of the 15q11.2 CNV, we also derived normative z-scores for cerebral brain features computed using FreeSurfer (Fischl, 2012). For these anatomical features, we derived z-scores using normative models based on data from 40k individuals and 58k individuals (Rutherford, Fraza, et al., 2022; Rutherford et al., 2023). Results largely replicated previous reports (Writing Committee for the ENIGMA-CNV Working Group et al., 2019) and gave effects sizes ranging from 0.34 to −0.29, i.e. comparable to the effects seen for cerebellar morphological features. See Supplementary Results and Supplementary Figure 4-5 for full results.

## Discussion

The current study, investigating both mean group differences and individual effects in cerebellar morphology in 15q11.2 CNV carriers yielded three main findings all characterized by small effect sizes. First, at the group level carriers of both deletions and duplications demonstrated smaller total cerebellar volumes relative to non-carriers. Secondly, a few specific regional differences emerged: 15q11.2 deletion carrier status was linked to smaller volume in the cerebellar right Crus I and Vermis VII, while 15q11.2 duplication carriers showed smaller volume in the right lobule IV and left lobule VI. Third, the investigation at the individual level uncovered heterogenous pattern among individuals in the carriers explaining the small effects observed.

The cerebellum undergoes a protracted development (Sathyanesan et al., 2019) where each region following a distinct developmental trajectory (Kim et al., 2024). As a result, certain cerebellar regions might be more vulnerable to risk of environmental and genetic disruptions including CNVs (Sefik et al., 2024). Cerebellar regions, including Vermis VIII-X and the cerebellar cortex, have been found to be particularly sensitive to CNVs (Modenato et al., 2021). Thus, we used normative modelling to identify the impact of the CNVs on both total and regional cerebellar morphology, by examining deviations and individual-level differences from the norm.

Aside from the 15q11.2 region being previously identified for its modest effect on brain structure (Sønderby et al., 2022), we observed small effects on the cerebellum. The case-control analysis revealed that 15q11.2 duplication carriers showed reduced volume in the right lobule IV and left VI, an area involved in various motor functions. Conversely, deletions of 15q11.2 exhibited reduced volume in the right Crus I and Vermis VII, regions associated with complex mentalizing events, person perception, and abstraction, including social action sequences (Buckner et al., 2011; Heleven et al., 2019; Schmahmann, 2010; Van Overwalle et al., 2020). Deletions at the 15q11.2 demonstrated a larger effect size than duplications in total and regional cerebellar volume, implying deletions may have more impact. Previous studies show developmental delay including motor, speech, language and cognitive skills were more prevalent in individuals with 15q11.2 deletions (Burnside et al., 2011; Cox & Butler, 2015; Stefansson et al., 2014). Reduced expression of the four genes situated in the 15q11.2 (BP1-BP2) deletion (Chai et al., 2003) may influence the cerebellum’s abnormal motor functions. Notably, *CYFIP1* has been linked to abnormalities in motor coordination in mice (Domínguez-Iturza et al., 2019), SZ (Nebel et al., 2016) and ASD (Wang et al., 2015). This is noteworthy, even considering the incomplete penetrance observed in non-affected individuals with CNV carriers, as it aligns with the motor and developmental delays associated with this deletion (Cafferkey et al., 2014).

Given the known reduced penetrance from 2 to 10.4% for deletions (Rosenfeld et al., 2013; Vassos et al., 2010) and the variable expressivity (Cox & Butler, 2015) associated with 15q11.2, there is likely to be considerable heterogeneity. Our examinations of individual-level extreme deviations in 15q11.2 CNV carriers mapped heterogeneous patterns of extreme positive and negative deviations among individuals with the same genetic variations, indicating a degree of heterogeneity. We observed that carriers of 15q11.2 deletions and duplications exhibited higher frequencies of extreme negative deviations in several ROIs compared to noncarriers, although no differences were observed in total cerebellar volume. Among the 28 ROIs, the maximum proportion of extreme deviations reached only to 8.3%, underscoring the absence of a single prominent area in extreme deviations. This finding shows that the effect of CNV on the cerebellar is multifaceted.

In addition to replicating previously reported cortical and subcortical results in a partially overlapping sample of 15q11.2 deletions and duplications (Writing Committee for the ENIGMA-CNV Working Group et al., 2019), we estimated the extreme deviations of CNVs in all global and subcortical volumes using normative modelling. Similarly, a recent study of the 1q21.1 distal CNV (Fraza et al., 2024) showed large negative deviations in the 1q21.1 duplications in the cerebellum, brainstem and pallidum. The overlap of findings concerning the cerebellum in these studies suggests it may be relevant to understanding the implications of carrying a pathogenic CNV.

This study has several limitations. First, the pleiotropic nature of CNVs complicates understanding their biological effects, as a single CNV can influence multiple phenotypic traits through diverse mechanisms, leading to heterogeneous clinical presentations and shared etiologies across neurodevelopmental and neuropsychiatric disorders (Consortium, 2019) (Jensen & Girirajan, 2017, 2019). This complexity challenges efforts to disentangle specific genotype-phenotype relationships and interpret morphological changes. Consistent with this, we observed highly individualized cerebellar effects among CNV carriers. CNV carriers also often display a broad range of overlapping cognitive, psychiatric, and physical traits (Mollon et al., 2023), further complicating the identification of consistent brain structural correlates. Small effect sizes suggest that while some carriers show cerebellar alterations, many do not, though broader medical and psychiatric risks remain (Crawford et al., 2019). Additionally, studying rare CNVs requires large, representative samples, which are difficult to obtain. This challenge is compounded by the reliance on UK Biobank data, which carries healthy volunteer bias and may limit generalizability (Farrell et al., 2020; Fry et al., 2017). Sample independence is another concern, as the normative models included data from the same cohort. While normative modeling is a valuable tool, overlapping samples may introduce subtle biases. Finally, despite advances in genetic and neuroimaging research, establishing causal pathways from genetic variation to brain morphology and clinical outcomes remains difficult due to the interplay of biological, environmental, and developmental factors (Andreassen et al., 2023; Cook Jr & Scherer, 2008).

## Conclusion

This cerebellar normative modeling study identified significant, though modest, differences in cerebellar morphology among 15q11.2 CNV carriers. Group-level analyses revealed smaller total and regional cerebellar volumes in both deletion and duplication carriers. However, individual-level deviation analyses highlighted considerable heterogeneity, with some carriers exhibiting pronounced morphological alterations while others did not. Notably, we observed no evidence of dosage effects across copy number variations. These findings suggest that CNVs at the 15q11.2 locus exert subtle and highly individualized influences on cerebellar structure. Our approach advances our understanding of the cerebellum’s role in the neurodevelopmental impacts of CNVs and underscores the importance of personalized assessments in genetic neuroimaging research.

## Methods

### Population Data

The individuals without diagnoses for normative modelling were obtained from the following studies: ABIDE, ADHD200, AOMIC ID1000, Beijing Enhanced, CAMCAN, CoRR, DLBS, DS000119, DS000202, DS000222, Fcon1000, HBN, HCP, MPI Lemon, NKI-Rockland, OASIS-3, PING, SALD, SLIM, and UK Biobank. Data used in the preparation of this article were obtained from the Alzheimer’s Disease Neuroimaging Initiative (ADNI) database (adni.loni.usc.edu). The ADNI was launched in 2003 as a public-private partnership, led by Principal Investigator Michael W. Weiner, MD. The primary goal of ADNI has been to test whether serial magnetic resonance imaging (MRI), positron emission tomography (PET), other biological markers, and clinical and neuropsychological assessment can be combined to measure the progression of MCI and early AD. Additional information about each study can be found in their respective publications (Supplementary Table 1). Table 1 summarizes the demographic information of individuals without diagnosis of 26509 (53% females) in training set and 26378 in testing set (53% female) which does not include individuals with CNV carriers. The age range spanned from 3 to 85 years. In this study, we included individuals with deletions and duplications at the 15q11.2 (n= 120 for deletions and n=149 duplication). The criteria for selecting CNVs required more than 100 participants for both deletions and duplications, thereby ensuring robust statistical power for the analyses. Individuals were excluded if baseline scans, demographic information, T1-weighted MRI data were missing or have withdrawn. In analyses when compared to non-carriers, individuals in the UK Biobank from the testing set were used. The global and subcortical regions were standardized using the standardized image analysis using FreeSurfer Software Suite version 5.1 to 5.3 (FreeSurfer). The CNV carrier datasets were obtained from the UK Biobank, where we only included CNVs of 15q11.2 deletions and duplications with outputs of ACAPUCLO and SUIT. We used the CNV calls including the quality control from the study of (Crawford et al., 2019) where, in brief, individuals were excluded if they had more than 30 CNVs, a waviness factor >0.03 or <−0.03, a call rate <96% or log R ratio SD >0.35.

**Table 1.**
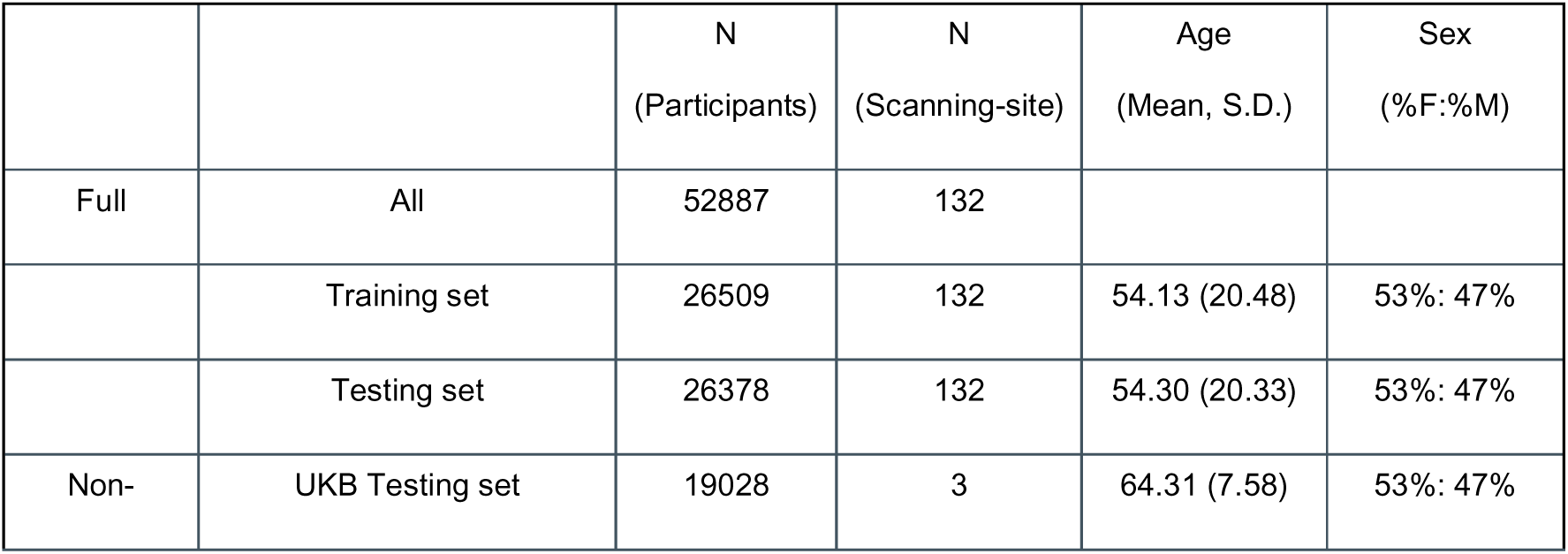
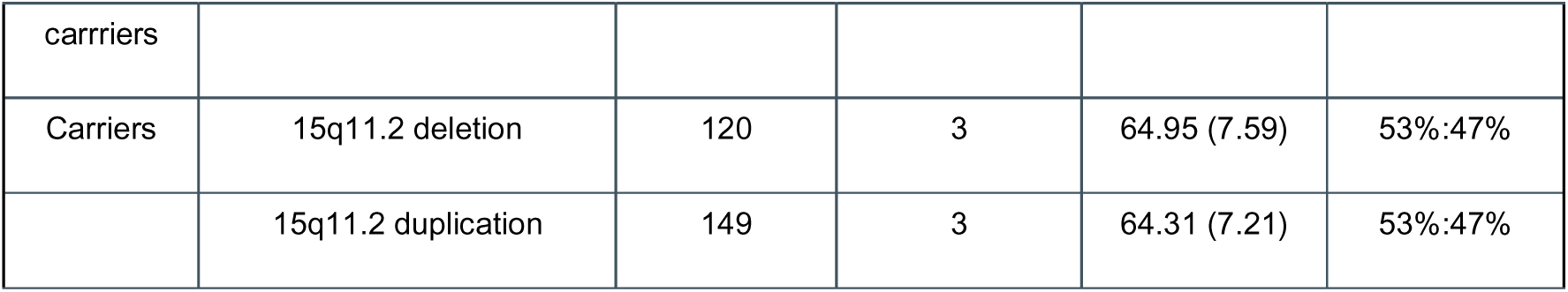
Sample description and demographics.

|  |  | N<br>(Participants) | N<br>(Scanning-site) | Age<br>(Mean, S.D.) | Sex<br>(%F:%M) |
| --- | --- | --- | --- | --- | --- |
| Full | All | 52887 | 132 |  |  |
|  | Training set | 26509 | 132 | 54.13 (20.48) | 53%: 47% |
|  | Testing set | 26378 | 132 | 54.30 (20.33) | 53%: 47% |
| Non- | UKB Testing set | 19028 | 3 | 64.31 (7.58) | 53%: 47% |
| carriers |  |  |  |  |  |
| Carriers | 15q11.2 deletion | 120 | 3 | 64.95 (7.59) | 53%:47% |
|  | 15q11.2 duplication | 149 | 3 | 64.31 (7.21) | 53%:47% |

### Lobular-level processing

As in our previous article (Kim et al., 2024), the T1-weighted images were skull stripped using the FreeSurfer 5.3 auto-recon pipeline (Ségonne et al., 2004) and reoriented to the standard FSL orientation using the *fslreorient2std* (Jenkinson et al., 2012). The linear registration was performed using *flirt* (Jenkinson & Smith, 2001), which utilized linear interpolation and the default 1 mm FSL template (version 6.0). The borders were cropped in [6:173, 2:214, 0:160] coordinates to minimize size while retaining all brain tissue. Lastly, the voxel intensity values were normalized to the range of [0,1], adjusting the intensity values of each voxel to a standardized scale.

As a part of ENIGMA Cerebellum Volumetric Pipeline, ACAPULCO algorithm (Han et al., 2020), a cerebellum parcellation algorithm based on convolutional neural networks, delivers fast and precise quantitative in-vivo regional assessment of the cerebellum. Instead of utilizing raw T1 images, we used pre-processed T1 images.

N4 (Tustison et al., 2010) was applied to correct Inhomogeneity. Moreover, the images were then registered to the 1mm isotropic ICBM 2009c template in MNI space using the ANTs registration suite (Fonov et al., 2011). The ACAPULCO algorithm is based 15 expert manual delineations of an adult cohort (Carass et al., 2018), achieves per-voxel labelling and employs post-processing for accurate segmentation. ACAPULCO segments cerebellum into 28 cerebellar lobules and computes the volume (mm^3^): bilateral Lobules I–VI; Crus I and II; Lobules VIIB, VIIIA, VIIIB, and IX-X; Vermis VI, VII, VIII, IX, and X; and Corpus Medullare (CM). Based on the automated quality control, certain participants with the extreme outliers whose number of lobules exceeded a certain threshold (n > 2) were removed.

### Voxel-level processing

In the same way as our previous study, we used the SUIT (Spatially Unbiased Infratentorial Toolbox) toolbox to segment cerebellar grey and white matter voxel-based morphometry (VBM) maps. This SUIT process leverages the outputs from ACAPULCO, MNI-aligned T1 image (Diedrichsen, 2006; Diedrichsen et al., 2009), and average mask derived from randomly selected 300 individuals without diagnosis. After segmentation, the grey matter maps are normalized for standard comparison and re-sliced to align in standardized space. Moreover, the grey matter maps were modulated by the Jacobian to preserve the value of each voxel in proportion to its original volume. Jacobian modulation was applied to proportionally maintain values of the original volume.

### Normative modeling

Figure 1B illustrates normative growth trajectories for cerebellar lobules that span from ages 3 to 85 (see Supplementary Fig 1 for all regions). The model performance is reported by explained variance, kurtosis, skew, and mean squared logarithmic loss (MSLL) which are shown in Supplementary Figure 2 and these are comparable to our previous studies (Kim et al., 2024) which included CNV carriers in the training set.

The current method is identical to the method in the cerebellar normative model described by Kim and colleagues (Kim et al., 2024), except that CNV carriers were excluded from the training set. We estimated normative model on cerebellar volume and voxel-wise intensity accounting for sex, age, and scanning site using PCNtoolkit package (version 0.34) in Python 3.8 (Marquand et al., 2019; Rutherford, Kia, et al., 2022). We used Bayesian Linear Regression (BLR) with likelihood warping approach (Rios & Tobar, 2019), with ‘sinarcsinh’ transformation (Dinga et al., 2021; Fraza et al., 2021) which provide evaluation metrics including explained variance, mean squared log-loss, skew, and kurtosis (Dinga et al., 2021). For surface area, cortical thickness, and subcortical volume, we used BLR normative models of existing surface area, cortical thickness, and subcortical volume (Rutherford et al., 2023; Rutherford, Fraza, et al., 2022).

### Analysis

We employed a one-way ANOVA to assess differences between groups concerning the CNVs. The independent variable was group (e.g., deletions, duplications, non-carriers), and the dependent variable was the volumetric measures of the cerebellar regions. Prior to performing ANOVA, we verified the assumptions of normality and homogeneity of variance using Shapiro-Wilk and Levene’s tests. Significant omnibus ANOVA results were followed up by independent samples t-tests to compare the deviation scores of deletions or duplications of CNVs with those of non-carriers. Additionally, we used nonparametric Mann-Whitney U-tests (Mann & Whitney, 1947) for the voxel-wise comparisons. For multiple comparison correction, we employed matrix spectral decomposition (Cheverud, 2001; Li & Ji, 2005; Nyholt, 2004) resulting in 19 independent features for the cerebellum. For global and subcortical volumes, we identified 10 independent features.

We calculated the percentage of extreme positive and negative deviations by counting the number of extreme deviations, defined as |z| > 1.96, and dividing this count by the total number of non-carriers or individuals with CNVs. To evaluate the frequencies of extreme deviations, we conducted chi-square tests of independence followed by post-hoc Z-tests to investigate the differences between groups. We also performed dose-response analyses, coding deletion carriers as 1, non-carriers as 2, and duplication carriers as 3.

## Supporting information

Supplementary Methods and Results

## Data availability

In this study, we used brain imaging from ABIDE, ADHD200, AOMIC ID1000, Beijing Enhanced, CAMCAN, CoRR, DLBS, DS000119, DS000202, DS000222, Fcon1000, HBN, HCP, MPI Lemon, NKI-Rockland, OASIS-3, PING, SALD, SLIM and UK Biobank, ADNI, AIBL, DEMGEN, PNC, and TOP. The cerebellar normative model is available on via PCNportal (Barkema et al., 2023): https://pcnportal.dccn.nl/.

## Code availability

Code for normative model is available as open-source python package, Predictive Clinical Neuroscience (PCN) toolkit (https://github.com/amarquand/PCNtoolkit). Further codes are available on http://github.com/milinkim/cerebellum_cnv.

## Ethics of the study

In this study, we used both publicly and privately available data. For detailed information regarding the consent and ethical procedures of each study, please refer to the Supplementary Information. All data were securely stored on the University of Oslo’s secure platform, Services for Sensitive Data (TSD), in compliance with Norwegian privacy regulations. The study was approved by the Regional Committee for Medical and Health Research Ethics, South-East Norway (REK, ref. no. 2019/943).

## Roles of funding

The work was supported by the South-Eastern Norway Regional Health Authority (2021040, supporting M.K. & T.M.; 2018037, 2018076, 2019101, 2021070, 2023012, 500189, #2020060 (supporting IES)), NIH R01MH129858 (IES), DFG Emmy Noether 513851350 and BMBF/DLR FEDORA 01EQ2403G (supporting T.W.), NordForsk (#164218), the Research Council of Norway (249795, 248238, 276082, 286838, 288083, 323951, 324499), Stiftelsen Kristian Gerhard Jebsen (SKGJ-MED-021), ERA-Net Cofund through the ERA PerMed project IMPLEMENT, and the European Research Council under the European Union’s Horizon 2020 research and Innovation program (ERC StG Grant No. 802998). We performed this work on the Services for sensitive data (TSD), University of Oslo, Norway, with resources provided by UNINETT Sigma2 - the National Infrastructure for High-Performance Computing and Data Storage in Norway. The funders had no role in the conception of the study, the analyses, or the interpretation of the results.

## Conflict of Interest Disclosures

O.A.A. has received speaker fees from Lundbeck, Janssen, Otsuka, and Sunovion and is a consultant to Cortechs.ai and Precision-Health.ai. E.H.L. has received speaker fees from Lundbeck, and is the CTO and shareholder of baba.vision. L.T.W. and T.W. are shareholders of baba.vision. KP contributed to clinical trials for Roche (BN29553) and Novo Nordisk (NN6535-4730), outside the submitted work. The other authors report no competing interests. G.S. has participated in Advisory Board meetings for Roche, Eli-Lilly, and Eisai regarding disease-modifying drugs for Alzheimer’s disease and has received honoraria for delivering lectures at symposia sponsored by Eisai and Eli-Lilly.

## Acknowledgements

We are grateful to all the individuals who participated in the studies and acknowledge the contributions of the clinicians and researchers involved in the recruitment and assessment of participants for making this work possible. We conducted this research using the UK Biobank Resource under application number 27412.

Data collection and sharing for this project was funded by the Alzheimer’s Disease Neuroimaging Initiative (ADNI) (National Institutes of Health Grant U01 AG024904) and DOD ADNI (Department of Defense award number W81XWH-12-2-0012). ADNI is funded by the National Institute on Aging, the National Institute of Biomedical Imaging and Bioengineering, and through generous contributions from the following: AbbVie, Alzheimer’s Association; Alzheimer’s Drug Discovery Foundation; Araclon Biotech; BioClinica, Inc.; Biogen; Bristol-Myers Squibb Company; CereSpir, Inc.; Cogstate; Eisai Inc.; Elan Pharmaceuticals, Inc.; Eli Lilly and Company; EuroImmun; F. Hoffmann-La Roche Ltd and its affiliated company Genentech, Inc.; Fujirebio; GE Healthcare; IXICO Ltd.; Janssen Alzheimer Immunotherapy Research & Development, LLC.; Johnson & Johnson Pharmaceutical Research & Development LLC.; Lumosity; Lundbeck; Merck & Co., Inc.; Meso Scale Diagnostics, LLC.; NeuroRx Research; Neurotrack Technologies; Novartis Pharmaceuticals Corporation; Pfizer Inc.; Piramal Imaging; Servier; Takeda Pharmaceutical Company; and Transition Therapeutics. The Canadian Institutes of Health Research is providing funds to support ADNI clinical sites in Canada. Private sector contributions are facilitated by the Foundation for the National Institutes of Health (www.fnih.org). The grantee organization is the Northern California Institute for Research and Education, and the study is coordinated by the Alzheimer’s Therapeutic Research Institute at the University of Southern California. ADNI data are disseminated by the Laboratory for Neuro Imaging at the University of Southern California. Also, data used in preparation of this article were obtained from the Australian Imaging Biomarkers and Lifestyle Study of Ageing (AIBL) databases (adni.loni.usc.edu).

## Authorship Contributions

T.M., T.W., and M.K. originally conceived of the project. M.K., T.W., T.M., and E.H.L. performed the analyses. M.K. T.W. T.M. wrote the initial draft of the manuscript. O.A., G.R., K.P., G.S., N.E.S., O.B.S., D.A., T.W. and L.W. contributed to data curation. All authors discussed the results and contributed to the final manuscript.

## Notes

### Author Declarations

These are analyses of publicly and privately available data. Description of informed consent and other ethical procedures is extensively described in each study, referenced in the manuscript. The data were stored and analyzed using University of Oslos secure platform, Services for sensitive data (TSD), in compliance with Norwegian privacy regulations. The study was approved by the Regional Committee for Medical and Health Research Ethics, South-East Norway (REK, ref. no. 2019/943)

