## Supplementary Methods and Results for "Mapping Cerebellar Morphology in 15q11.2 CNV Carriers Using Normative Modeling"

***Table of Contents***

**Kim et al.**

**Supplementary Methods**

**Supplementary Results**

**List of Supplementary Figures**

1. Cerebellar lobular growth charts
2. Evaluation metric of the cerebellar growth charts
3. Overlap of extreme deviations in voxel-wise
4. The Association with global and subcortical brain morphology

**List of Supplementary Tables**

1. Sources of the studies used in the study
2. ANOVA summary of CNVs in ROI
3. Associations of CNVs with cerebellar ROI
4. Percentage of Extreme deviations in ROI of CNVs
5. Chi square test
6. Posthoc test of Z-test
7. Dose effect of CNVs
8. Percentage of Extreme deviations in Subcortical of CNVs
9. Percentage of Extreme deviations in Surface Area of CNVs
10. ANOVA summary of CNVs in Subcortical
11. Associations of 15q11.2 with Global and Subcortical Brain Morphology

**Supplementary Methods**

***Statistics & Reproducibility***

The quality control of this study is identical the procesude from Kim and colleagues^1^ utilizing the full sample of the cerebellar normative model. We perfromed quality control by running the ENIGMA Cerebellum Volumetric Pipeline QC Scripts of ACAPULCO^2^ with Singularity^3^. Given the extensive volume of brain scans available, amounting to thousands, it was not feasible to manually inspect each individual scan. However, approximately 5% of these scans were selected and examined manually to ensure quality and consistency within the dataset. The “QC_Images.html” file displays visual representations of the segmented images in coronal, sagittal, and transverse sections, which allows for a thorough inspection of the segmentation quality. The QC pipeline delivers both quantitative and visual information aids pertaining to the volumetric aspects of the cerebellum's segmented regions including volume, outliers, and box plots of the outliers. By integrating quality control pipeline into our analysis, it enabled us to detect and rectify any mis-segmentations or statistical outliers, thereby enhancing the dependability and precision of our findings. We further excluded participants whose scans revealed outliers in at least two regions, as well as when a scanning site contributed data from fewer than five participants.

**Supplementary Results**

ANOVAs conducted on subcortical regions showed significant group effects in multiple areas (see Supplementary Table 10). 15q11.2 deletion carrier status was significantly associated with increased mean z-scores for cortical thickness (T=3.95, uncorrected *p* =0.0001, Cohen’s d = 0.34) and nucleus accumbens volume (T= -3.42, uncorrected *p* =0.0008, Cohen’s d = -0.28) compared to non-carriers, while duplication carriers showed smaller surface area compared to non-carriers (T=-3.21, uncorrected *p* =0.0016, Cohen’s d = -0.29) and intracranial volume (T= -2.98, uncorrected *p*=0.0034, Cohen’s d= -0.24) (Supplementary Figure 4A and Supplementary Table 10-11). The percentages of extreme negative deviations of global and subcortical morphology are shown in Figure 4B and Supplementary Table 8-9.

**Supplementary Figures**

| 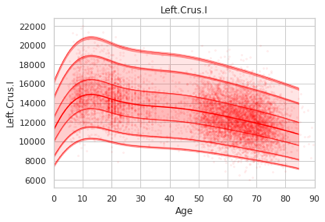 | 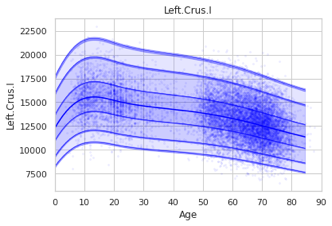 | 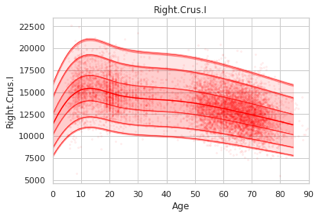 | 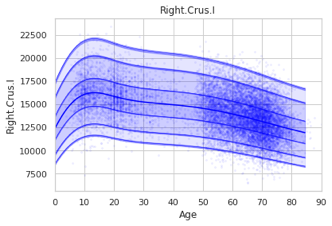 |
| --- | --- | --- | --- |
| 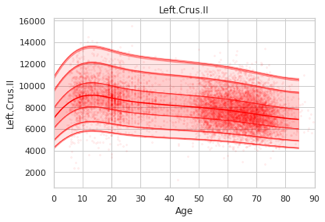 | 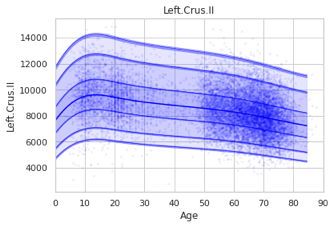 | 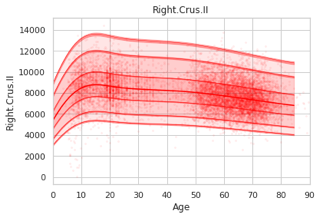 | 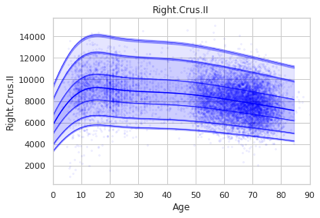 |
| 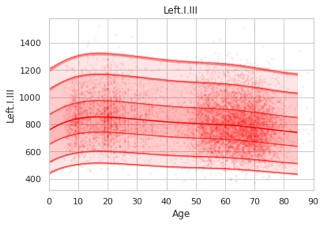 | 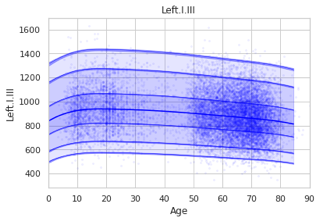 | 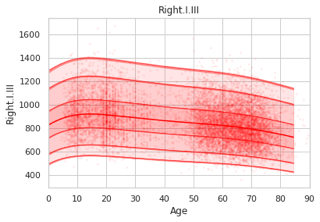 | 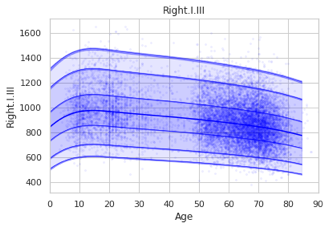 |
| 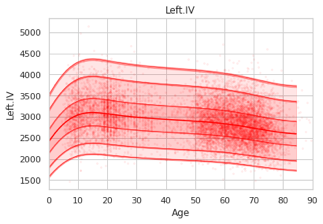 | 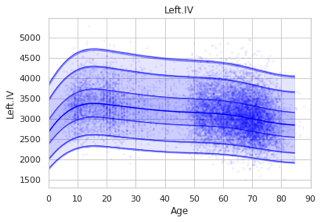 | 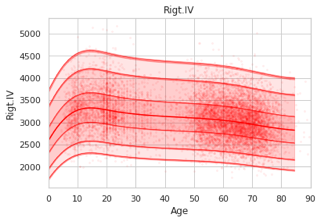 | 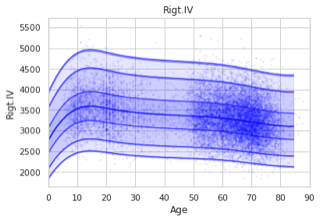 |
| 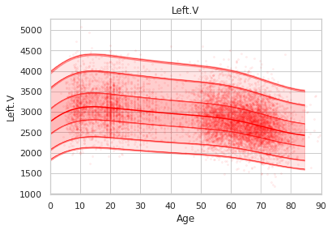 | 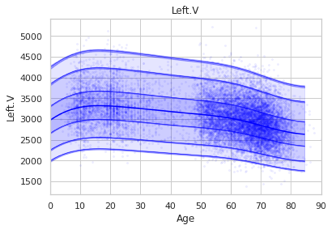 | 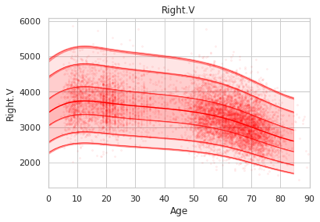 | 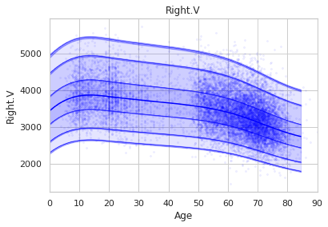 |
| 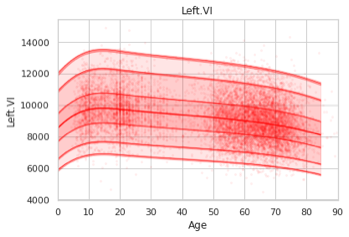 | 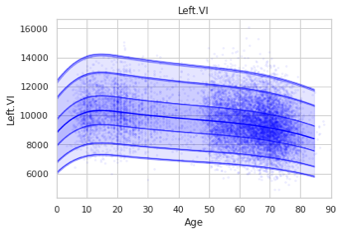 | 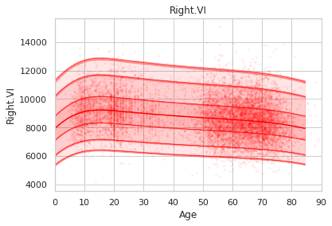 | 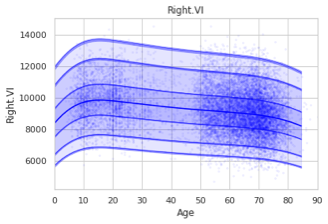 |
| 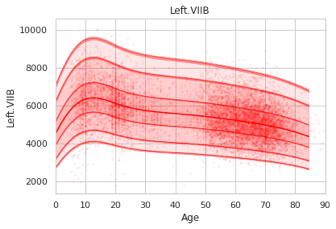 | 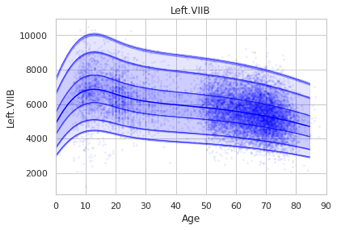 | 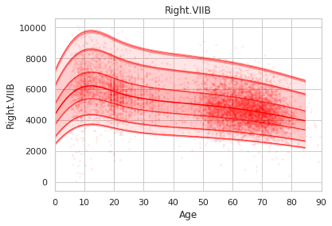 | 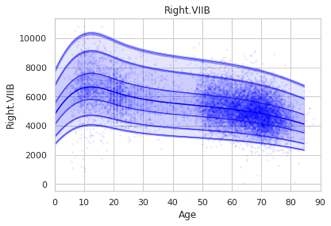 |
| 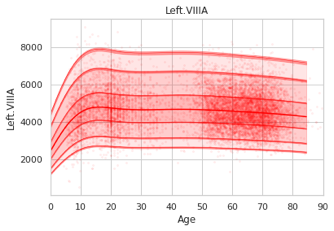 | 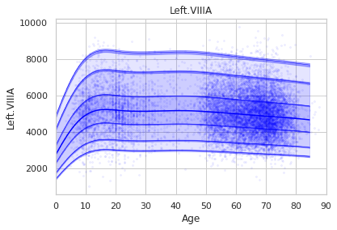 | 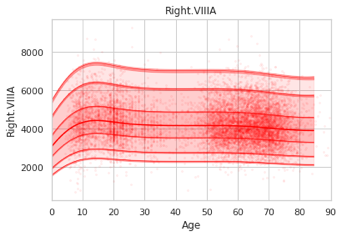 | 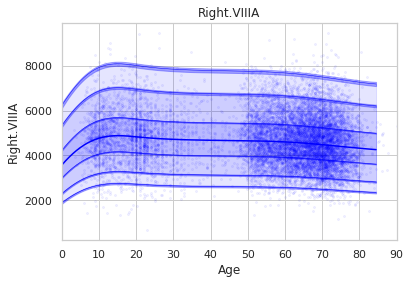 |
| 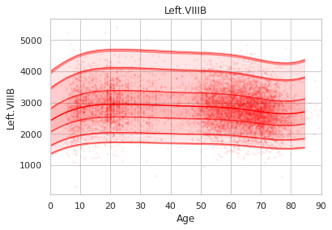 | 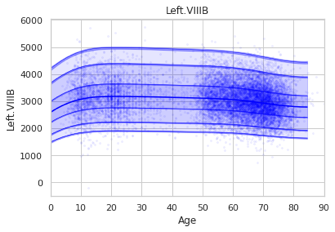 | 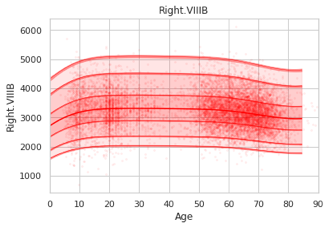 | 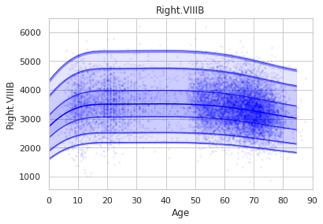 |
| 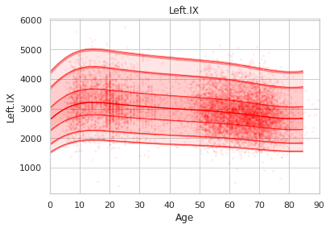 | 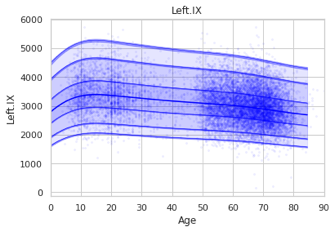 | 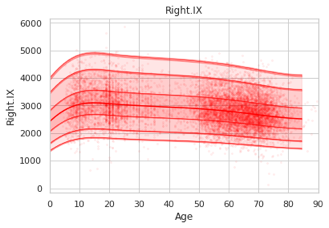 | 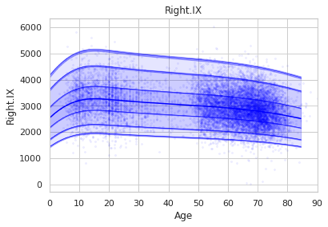 |
| 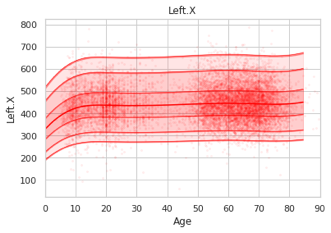 | 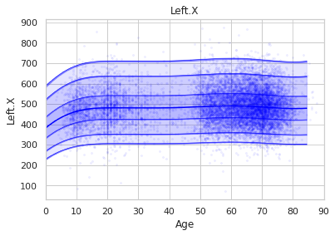 | 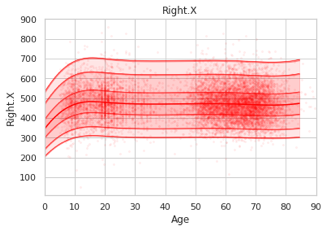 | 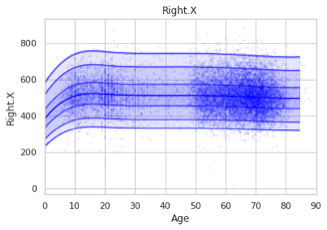 |
| 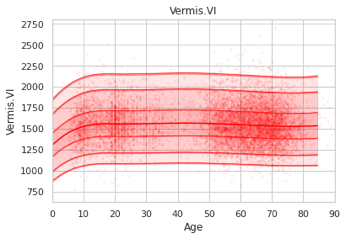 | 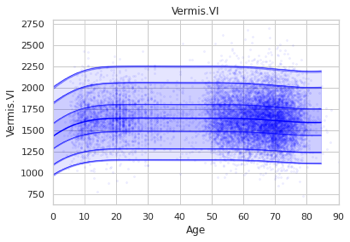 | 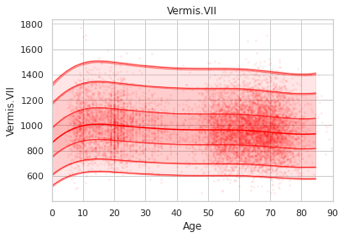 | 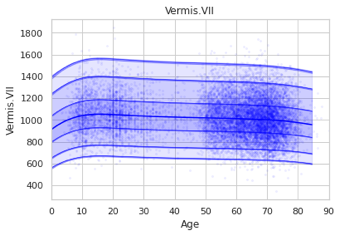 |
| 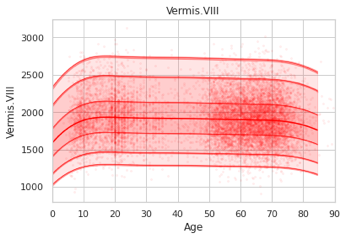 | 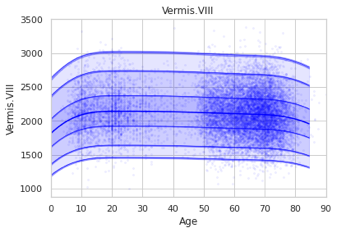 | 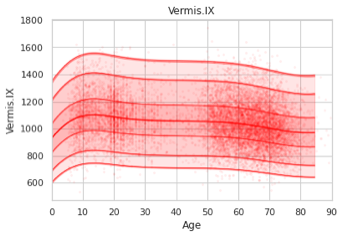 | 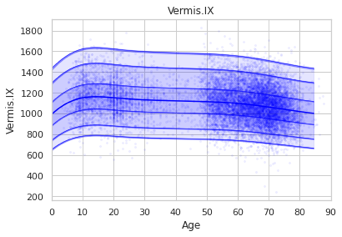 |
| 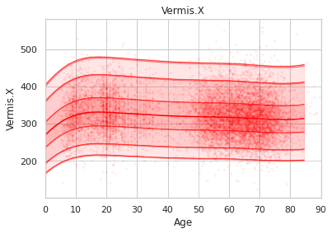 | 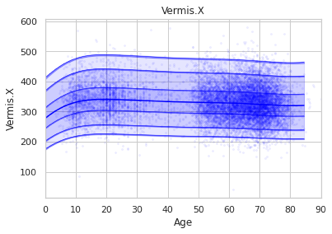 | 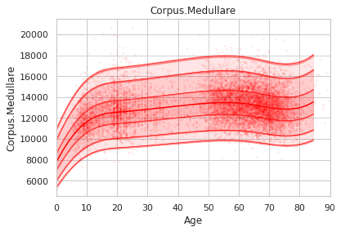 | 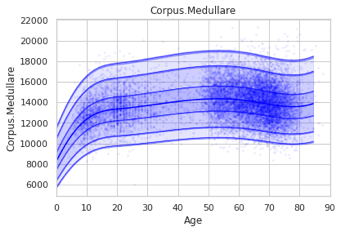 |
| 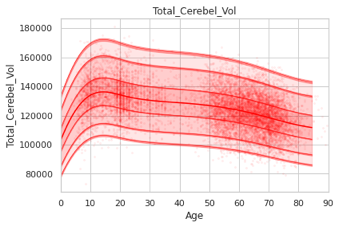 | 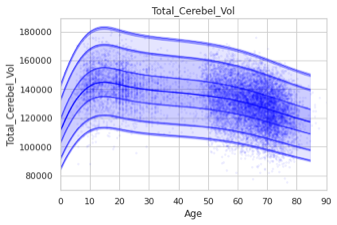 |  |  |

**Supplementary Figure 1. Cerebellar lobular growth charts.** Total of 28 anatomical regions of the cerebellum are mapped throughout the lifespan, including total cerebellum volume. This is re-estimated without CNV-carriers in the training set. The red figure is female and blue figure is male.

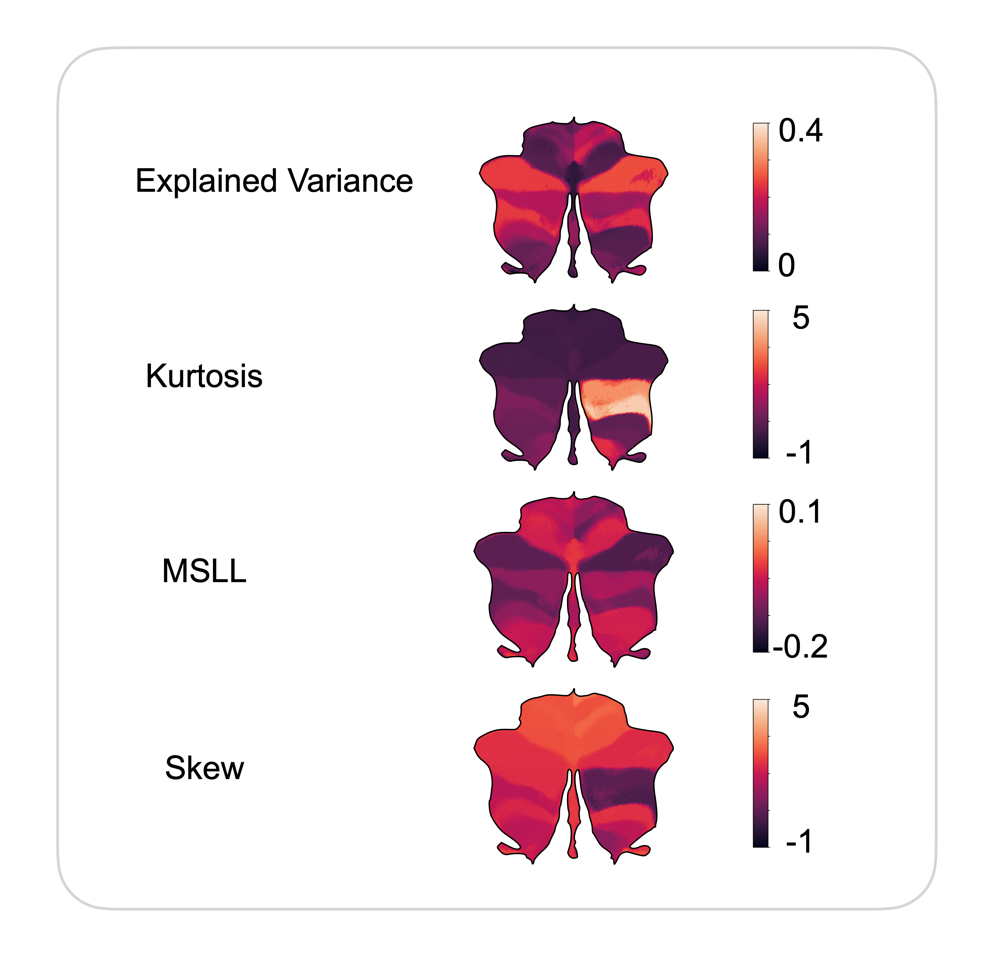

**Supplementary Figure 2. Evaluation metric of the cerebellar growth charts.** Evaluation metrics across test set show a good fit of the model in central tendency and variance (explained variance, kurtosis) and shape of the model (MSLL and skew).

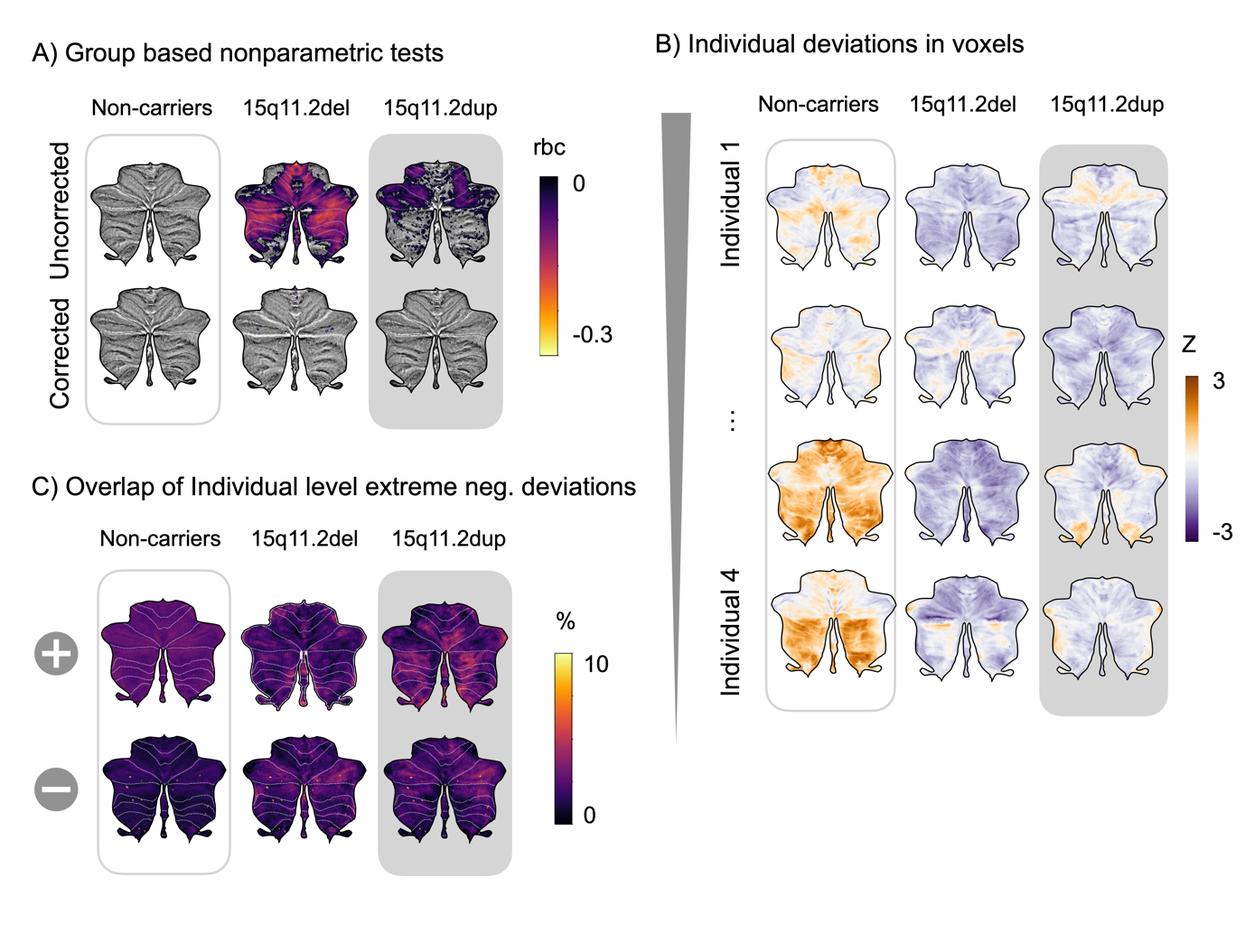

**Supplementary Figure 3. Overlap of extreme deviations in voxel-wise of 15q11.2.** (A) Individual extreme positive and negative deviations for non-carriers and CNVs show unique patterns across the cerebellum. (B) The scale shows overlap of positive and negative extreme deviations in each cohort in voxels. (C) The scale shows the two-sided Mann-Whitney U-test rank biserial correlation (rbc) for individual CNVs against non-carriers (NC), with results presented both uncorrected and corrected for multiple comparisons, highlighting negative effects.

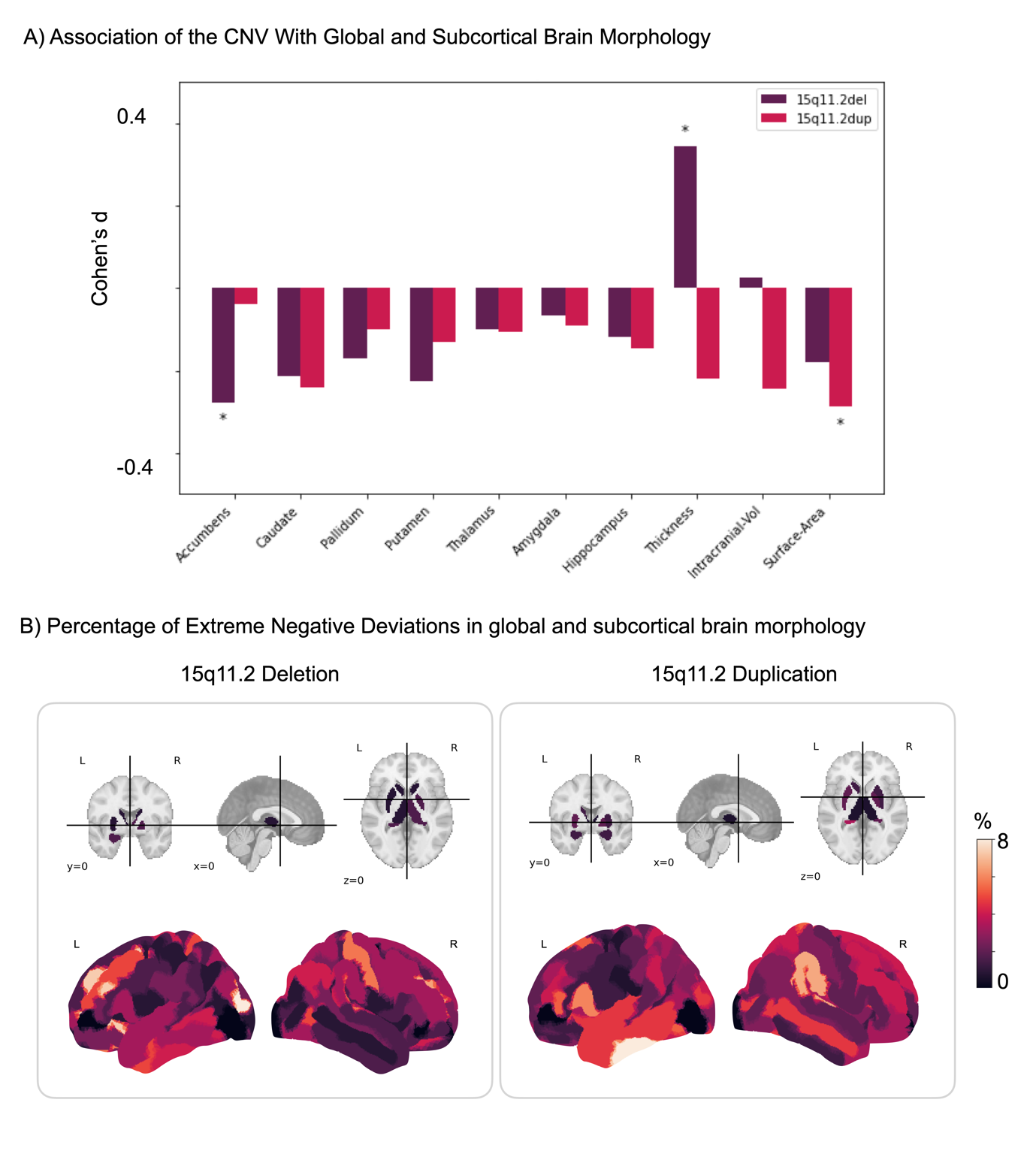

**Supplementary Figure. 4. The 15q11.2 CNVs are associated with global and subcortical brain morphology.** (A) Compared to non-carriers, individuals with 15q11.2 deletions show increased cortical thickness but a reduced size of the accumbens. In contrast, duplications are associated with a decrease in surface area. These findings have been adjusted for multiple comparisons**.** (B) The analysis of extreme negative deviations in subcortical regions reveals that for 15q11.2 duplications, the most significant deviations are observed in the cerebellum cortex and hippocampus. In contrast, for deletions, the primary region exhibiting these negative deviations is the cerebellum. When considering surface area, the right superior transverse temporal region shows the highest deviations associated with duplications, while the left superior occipital middle region and lunate sulcus demonstrate the greatest deviations for deletions.

**Supplementary Tables**

**Supplementary Table 1. Sources of the studies used in the study**

| **Datasets** | **Sources** | **Comments** | **References** |
| --- | --- | --- | --- |
| Autism Brain  Imaging Dataset  Exchange | http://fcon_1000.projects.nitrc.org/ | Primary support for the work by Adriana Di Martino was provided by the NIMH  (K23MH087770) and the Leon Levy Foundation. Primary support for the work by Michael P. Milham and the INDI team was provided by gifts from Joseph P. Healy and the Stavros Niarchos Foundation to the Child Mind Institute, as well as by an NIMH award to MPM (R03MH096321). | ^4^ |
| Autism Brain  Imaging Dataset  Exchange II | http://fcon_1000.projects.nitrc.org/ | Primary support for the work by Adriana Di Martino and her team was provided by the National Institute of Mental Health (NIMH 5R21MH107045). Primary support for the work by Michael P. Milham and his team provided by the National Institute of Mental Health (NIMH 5R21MH107045); Nathan S. Kline Institute of Psychiatric Research). Additional Support was provided by gifts from Joseph P. Healey, Phyllis Green and Randolph Cowen to the Child Mind Institute. | ^5^ |
| ADHD200 | http://fcon_1000.projects.nitrc.org/ | F. Xavier Castellanos, David Kennedy, Michael Milham, and Stewart Mostofsky are responsible for the initial conception of the ADHD-200 Consortium. Consortium steering committee includes Jan Buitelaar, F. Xavier Castellanos, Dan Dickstein, Damien Fair, David Kennedy, Beatriz Luna, Michael Milham (Project Coordinator), Stewart Mostofsky, and Julie Schweitzer. Data aggregation and organization was coordinated by the INDI team, which included Saroja Bangaru, David Gutman, Maarten Mennes, and Michael Milham. Web infrastructure and data storage were coordinated by Robert Buccigrossi, Albert Crowley, Christian Hasselgrove, David Kennedy, Kimberly Pohland, and Nina Preuss. The ADHD-200 Global Competition Coordinators were Damien Fair (Chair of Selection Committee, Editor in Chief for Global Competition Special issue) and Michael Milham | ^6,7^ |
| Alzheimer’s  Disease  Neuroimaging  Initiative (ADNI) | <http://adni.loni.usc.edu/> | The ADNI was launched in 2003 as a public-private partnership, led by Principal Investigator Michael W. Weiner, MD. ADNI consists of 4 waves, the later is still ongoing (ADNI 3). A complete listing of ADNI investigators can be found at  http://adni.loni.usc.edu/wpcontent/uploads/how_to_apply/ADNI_Acknowledgement_List.pdf. Data collection and sharing for this project was funded by the Alzheimer's Disease Neuroimaging Initiative (ADNI) (National Institutes of Health Grant U01 AG024904) and DOD ADNI (Department of Defense award number W81XWH-12-2-0012). ADNI is funded by the National Institute on Aging, the National Institute of Biomedical Imaging and Bioengineering, and through generous contributions from the following: AbbVie, Alzheimer's Association; Alzheimer's Drug Discovery Foundation; Araclon Biotech; BioClinica, Inc.; Biogen; Bristol-Myers Squibb Company; CereSpir, Inc.; Cogstate; Eisai Inc.; Elan Pharmaceuticals, Inc.; Eli Lilly and Company; EuroImmun; F. Hoffmann-La Roche Ltd and its affiliated company Genentech, Inc.; Fujirebio; GE Healthcare; IXICO Ltd.;Janssen Alzheimer Immunotherapy Research & Development, LLC.; Johnson & Johnson Pharmaceutical Research & Development LLC.; Lumosity; Lundbeck; Merck & Co., Inc.;Meso Scale Diagnostics, LLC.; NeuroRx Research; Neurotrack Technologies; Novartis Pharmaceuticals Corporation; Pfizer Inc.; Piramal Imaging; Servier; Takeda Pharmaceutical Company; and Transition Therapeutics. The Canadian Institutes of Health Research is providing funds to support ADNI clinical sites in Canada. Private sector contributions are facilitated by the Foundation for the National Institutes of Health (www.fnih.org). The grantee organization is the Northern California Institute for Research and Education, and the study is coordinated by the Alzheimer's Therapeutic Research Institute at the University of Southern California. ADNI data are disseminated by the Laboratory for Neuro Imaging at the University of Southern California. |  |
| The Australian  Imaging,  Biomarkers and  Lifestyle Flagship  Study (AIBL) | <https://aibl.csiro.au/> | Australian Imaging Biomarkers and Lifestyle flagship study of ageing (AIBL) was funded by the Commonwealth Scientific and Industrial Research Organisation (CSIRO), which was made available at the ADNI databas (http://www.loni.usc.edu/ADNI). The AIBL researchers contributed data but did not participate in analysis or writing of this report. AIBL researchers are listed at  http://www.aibl.csiro.au. Correspondence should be addressed to Christopher Rowe. | ^8^ |
| The Amsterdam  Open MRI  Collection (AOMIC) | https://nilab-uva.github.io/AOMIC.github.io/ | We thank all research assistants and students who helped collecting the data of the three projects, Jasper Wijnen and Marco Teunisse for advice and guidance with respect to anonymization and GDPR-related concerns, and Jos Bloemers, Sennay Ghebeab, Adriaan Tuiten, Joram van Driel, Christian Olivers, Ilja Sligte, Sara Jahfari, Guido van Wingen, and Suzanne Oosterwijk for help with designing the paradigms, Marcus Spaan for technical support, and Franklin Feingold and Joe Wexler for help with uploading the datasets to Openneuro. | ^9^ |
| Beijing Normal  University  Enhanced Sample (Bejing) | http://fcon_1000.projects.nitrc.org/ | Financial support for the data used in this project was provided by a grant from the National Natural Science Foundation of China: 30770594 and a grant from the National High Technology Program of China (863): 2008AA02Z405. | ^19,2019,20^ |
| Cam-CAN | https://camcan-archive.mrc-cbu.cam.ac.uk/dataaccess/ | Data collection and sharing for this project was provided by the Cambridge Centre for Ageing and Neuroscience (CamCAN). CamCAN funding was provided by the UK Biotechnology and Biological Sciences Research Council (grant number BB/H008217/1), together with support from the UK Medical Research Council and University of Cambridge, UK. | ^12,13^ |
| Consortium for  Reliability and  Reproducibility (Corr) | http://fcon_1000.projects.nitrc.org/ |  | ^14^ |
| Norwegian Dementia Genetics Network (DemGen) | Authors | Supported by the Norwegian National Advisory Unit on Aging and Health | ^15,16^ |
| Dallas Lifespan  Brain Study (fcon) | http://fcon_1000.projects.nitrc.org/ |  | ^17^ |
| ds00119 | https://openfmri.org/ | Supported by the National Institutes of Mental Health (NIMH RO1 MH067924). Enami Yasui  provided assistance with data collection | ^18^ |
| ds000202 | https://openfmri.org/ |  |  |
| ds000222 | https://openfmri.org/ |  | ^21^ |
| 1000 Functional  Connectomes  Classic Sample (fcon) | http://fcon_1000.projects.nitrc.org/ | Collected at 33 independent sites by J.J. Pekar, S.H. Mostofsky, S, Colcombe, Y.F. Zang, D. Marguiles, R.L. Buckner, M.J Low, B. Rypma, D.J. Madden, A.C. Evans, S.A.R.B. Rombouts, A. Villringer, S.J. Li, C.Sorg, V. Riedel, B. Biswal, M. Hampson, M.P. Milham, F.X. Castellanos, P. Williamson, M. Hoptman, V.J. Kiviniemi, J. Veijiola, S.M. Smith, C. Mackay, M. Greicius, G. Siegle, K. McMahon, B. Schlaggar, S. Petersen, C.P. Lin, H.S. Mayberg, C.S.  Monk, R.D. Seidler, S.J. Peltier |  |
| Human Connectome Project (HCP) | https://www.humanconnectome.org/ | Data were provided [in part] by the Human Connectome Project, MGH-USC Consortium (PIs: Bruce R. Rosen, Arthur W. Toga and Van Wedeen; U01MH093765) funded by the NIH Blueprint Initiative for Neuroscience Research grant; the National Institutes of Health grant P41EB015896; and the Instrumentation Grants S10RR023043, 1S10RR023401, 1S10RR019307. | ^22^ |
| Healthy Brain Network (HBN) | http://fcon_1000.projects.nitrc.org/ |  | ^23^ |
| Max Planck  Institut Leipzig  Mind-Brain-Body  Dataset (MPI) | http://fcon_1000.projects.nitrc.org/ |  | ^24^ |
| Enhanced Nathan  Kline Institute -  Rockland Sample (NKI) | http://fcon_1000.projects.nitrc.org/ | Principal support for the enhanced NKI-RS project is provided by the NIMH BRAINS R01MH094639-01 (PI Milham). Funding for key personnel also provided in part by the New York State Office of Mental Health and Research Foundation for Mental Hygiene. Funding for the decompression and augmentation of administrative and phenotypic protocols provided by a grant from the Child Mind Institute (1FDN2012-1). Additional personnel support provided by  the Center for the Developing Brain at the Child Mind Institute, as well as NIMH R01MH081218, R01MH083246, and R21MH084126. Project support also provided by the NKI Center for Advanced Brain Imaging (CABI), the Brain Research Foundation, and the Stavros Niarchos Foundation. | ^25^ |
| Open Access  Series of Imaging  Studies 3 (OASIS) | <http://www.oasis-brains.org/> | Data were provided by OASIS 3: Longitudinal Multimodal Neuroimaging: Principal Investigators: T. Benzinger, D. Marcus, J. Morris; NIH P30 AG066444, P50 AG00561, P30 NS09857781, P01 AG026276, P01 AG003991, R01 AG043434, UL1 TR000448, R01 EB009352. AV-45 doses were provided by Avid Radiopharmaceuticals, a wholly owned subsidiary of Eli Lilly. Supported by grants P50 AG05681, P01 AG03991, R01 AG021910, P50 MH071616, U24 RR021382, R01 MH56584. | ^26^ |
| Pediatric  Imaging,  Neurocognition  and Genetics (PING) | <http://pingstudy.ucsd.edu/> | Data used in the preparation of this article were obtained from the Pediatric Imaging,  Neurocognition and Genetics (PING) Study database (www.chd.ucsd.edu/research/ping-study.html,  now shared through the NIMH Data Archive (NDA)). PING was a multisite, cross-sectional  study that recruited more than 1,700 participants aged 3 to 20 years. The study was supported  by award number RC2DA029475 from the National Institute on Drug Abuse with additional  support for data sharing provided by the Eunice Kennedy Shriver National Institute of Child  Health & Human Development under award number R01HD061414. A list of participating sites  and study investigators can be found at https://ping-dataportal.ucsd.edu/sharing/Authors10222012.pdf.  PING investigators designed and implemented the study and/or provided data but did not  necessarily participate in analysis or writing of this report. This publication is solely the  responsibility of the authors and does not necessarily represent the views of the National  Institutes of Health or PING investigators. | ^27^ |
| Philadelphia Neurodevelopmental Cohort (PNC) | <https://www.med.upenn.edu/bbl/philadelphianeurodevelopmentalcohort.html> |  | ^28^ |
| Southwest  University Adult  Lifespan Dataset (SALD) | http://fcon_1000.projects.nitrc.org/ |  | ^14^ |
| Southwest University  Longitudinal  Imaging Multimodal Brain Data (SLIM) | http://fcon_1000.projects.nitrc.org/ | Support was provided by grant numbers 31271087; 31470981; 31571137, 31500885,  SWU1509383, SWU1509451, cstc2015jcyjA10106, 151023, 2015M572423, 2015M580767,  Xm2015037, 14JJD880009 | ^29,30^ |
| StrokeMRI (TOP) | Authors | Supported by the Research Council of Norway (249795, 248238), the South-Eastern Norway  Regional Health Authority (2014097, 2015044, 2015073, 2016083), and the Norwegian ExtraFoundation for Health and Rehabilitation (2015/FO5146) | ^31^ |
| Thematically Organized Psychosis (TOP) | Authors | Supported by several grants from the Research Council of Norway, and the South-Eastern  Norway Regional Health Authority | ^32–34^ |
| UK BioBank(UKB) | https://www.ukbiobank.ac.uk/ | This research has been conducted using the UK Biobank Resource (access code 27412). | ^35,36^ |

**Supplementary Table 2**. **ANOVA Summary of CNVs in ROIs**

The p-values that remain significant after adjustments for multiple comparisons are highlighted in bold.

| **ROI** | **F-statistic** | **ANOVA P-value** | **Eta Squared** | **Levene Statistic** | **Levene P-value** |
| --- | --- | --- | --- | --- | --- |
| Corpus.Medullare | 3.7345 | 0.0239 | 0.0004 | 0.1732 | 0.8409 |
| Left.Crus.I | 5.0927 | 0.0061 | 0.0005 | 0.0377 | 0.9630 |
| Left.Crus.II | 2.1033 | 0.1221 | 0.0002 | 3.0478 | 0.0475 |
| Left.I.III | 4.0592 | 0.0173 | 0.0004 | 0.3606 | 0.6973 |
| Left.IV | 0.4797 | 0.6190 | 0.0000 | 0.4393 | 0.6445 |
| Left.IX | 2.5313 | 0.0796 | 0.0003 | 3.5329 | 0.0292 |
| Left.V | 3.2322 | 0.0395 | 0.0003 | 0.0268 | 0.9736 |
| [**Left.VI**](http://Left.VI) | **7.1134** | **0.0008** | **0.0007** | **0.4211** | **0.6563** |
| Left.VIIB | 3.1940 | 0.0410 | 0.0003 | 0.1337 | 0.8749 |
| Left.VIIIA | 4.5486 | 0.0106 | 0.0005 | 0.8537 | 0.4259 |
| Left.VIIIB | 2.3783 | 0.0927 | 0.0002 | 1.1463 | 0.3178 |
| Left.X | 1.1219 | 0.3257 | 0.0001 | 1.9639 | 0.1403 |
| **Right.Crus.I** | **8.3717** | **0.0002** | **0.0009** | **0.1292** | **0.8788** |
| Right.Crus.II | 5.4455 | 0.0043 | 0.0006 | 2.5259 | 0.0800 |
| Right.I.III | 5.8289 | 0.0029 | 0.0006 | 0.2010 | 0.8180 |
| Right.IX | 1.0595 | 0.3467 | 0.0001 | 1.5008 | 0.2230 |
| Right.V | 1.3735 | 0.2532 | 0.0001 | 1.3609 | 0.2565 |
| [Right.VI](http://Right.VI) | 3.9973 | 0.0184 | 0.0004 | 0.7216 | 0.4860 |
| Right.VIIB | 2.9800 | 0.0508 | 0.0003 | 0.0790 | 0.9240 |
| Right.VIIIA | 0.2174 | 0.8046 | 0.0000 | 0.5889 | 0.5550 |
| Right.VIIIB | 4.5231 | 0.0109 | 0.0005 | 1.1621 | 0.3129 |
| Right.X | 3.0220 | 0.0487 | 0.0003 | 2.2017 | 0.1106 |
| **Rigt.IV** | **9.8000** | **0.0001** | **0.0010** | **0.0763** | **0.9266** |
| Vermis.IX | 2.1499 | 0.1165 | 0.0002 | 0.0604 | 0.9414 |
| [Vermis.VI](http://Vermis.VI) | 1.4180 | 0.2422 | 0.0001 | 1.6372 | 0.1946 |
| Vermis.VII | 5.2706 | 0.0051 | 0.0005 | 0.6832 | 0.5050 |
| Vermis.VIII | 1.0395 | 0.3537 | 0.0001 | 0.9494 | 0.3870 |
| Vermis.X | 0.9028 | 0.4055 | 0.0001 | 2.1466 | 0.1169 |
| **Total_Cerebel_Vol** | **7.3450** | **0.0006** | **0.0008** | **0.7544** | **0.4703** |

**Supplementary Table 3**. **Post-hoc Associations of CNVs with cerebellar ROI**

The p-values that remain significant after adjustments for multiple comparisons are highlighted in bold.

| **cnv** | **roi** | **t-stat** | **p-value** | **cohens d** | **median_hc** | **median_c** |
| --- | --- | --- | --- | --- | --- | --- |
| 15q11.2dup | Corpus.Medullare | -0.087 | 0.931 | -0.007 | 0.025 | 0.071 |
| 15q11.2dup | Left.Crus.I | -1.827 | 0.070 | -0.158 | 0.011 | -0.155 |
| 15q11.2dup | Left.Crus.II | -0.725 | 0.470 | -0.068 | 0.019 | 0.004 |
| 15q11.2dup | Left.I.III | -1.480 | 0.141 | -0.121 | 0.001 | -0.100 |
| 15q11.2dup | Left.IV | -0.444 | 0.658 | -0.038 | -0.001 | -0.049 |
| 15q11.2dup | Left.IX | -1.487 | 0.139 | -0.142 | 0.042 | -0.105 |
| 15q11.2dup | Left.V | -1.738 | 0.084 | -0.141 | 0.018 | -0.188 |
| **15q11.2dup** | **Left.VI** | **-3.121** | **0.002** | **-0.257** | **0.007** | **-0.305** |
| 15q11.2dup | Left.VIIB | -2.277 | 0.024 | -0.182 | 0.027 | -0.017 |
| 15q11.2dup | Left.VIIIA | -0.712 | 0.478 | -0.055 | 0.009 | 0.035 |
| 15q11.2dup | Left.VIIIB | -0.069 | 0.945 | -0.006 | 0.026 | 0.139 |
| 15q11.2dup | Left.X | 0.002 | 0.998 | 0.000 | -0.004 | -0.005 |
| 15q11.2dup | Right.Crus.I | -1.627 | 0.106 | -0.146 | 0.001 | -0.068 |
| 15q11.2dup | Right.Crus.II | -1.518 | 0.131 | -0.148 | 0.030 | -0.107 |
| 15q11.2dup | Right.I.III | -2.764 | 0.006 | -0.215 | -0.013 | -0.213 |
| 15q11.2dup | Right.IX | -1.049 | 0.296 | -0.094 | 0.039 | -0.167 |
| 15q11.2dup | Right.V | -1.464 | 0.145 | -0.121 | 0.006 | -0.118 |
| 15q11.2dup | Right.VI | -2.266 | 0.025 | -0.180 | 0.022 | -0.123 |
| 15q11.2dup | Right.VIIB | -1.957 | 0.052 | -0.169 | 0.014 | -0.105 |
| 15q11.2dup | Right.VIIIA | 0.712 | 0.477 | 0.054 | 0.004 | 0.055 |
| 15q11.2dup | Right.VIIIB | -0.535 | 0.594 | -0.047 | 0.021 | -0.053 |
| 15q11.2dup | Right.X | -1.478 | 0.141 | -0.132 | -0.021 | -0.092 |
| **15q11.2dup** | **Rigt.IV** | **-3.786** | **0.000** | **-0.302** | **0.000** | **-0.226** |
| 15q11.2dup | Vermis.IX | 0.337 | 0.737 | 0.028 | 0.011 | -0.029 |
| 15q11.2dup | Vermis.VI | 0.156 | 0.876 | 0.015 | -0.002 | -0.038 |
| 15q11.2dup | Vermis.VII | 0.109 | 0.913 | 0.010 | 0.024 | 0.109 |
| 15q11.2dup | Vermis.VIII | 0.346 | 0.730 | 0.029 | 0.025 | 0.034 |
| 15q11.2dup | Vermis.X | 1.391 | 0.166 | 0.113 | 0.017 | 0.146 |
| **15q11.2dup** | **Total Cerebellar V** | **-2.232** | **0.027** | **-0.205** | **0.050** | **-0.184** |
| 15q11.2del | Corpus.Medullare | -2.642 | 0.009 | -0.244 | 0.025 | -0.171 |
| 15q11.2del | Left.Crus.I | -2.391 | 0.018 | -0.221 | 0.011 | -0.098 |
| 15q11.2del | Left.Crus.II | -1.716 | 0.089 | -0.164 | 0.019 | -0.191 |
| 15q11.2del | Left.I.III | 2.178 | 0.031 | 0.189 | 0.001 | 0.143 |
| 15q11.2del | Left.IV | -0.579 | 0.564 | -0.055 | -0.001 | -0.215 |
| 15q11.2del | Left.IX | -1.130 | 0.260 | -0.098 | 0.042 | -0.133 |
| 15q11.2del | Left.V | -1.729 | 0.086 | -0.159 | 0.018 | -0.210 |
| 15q11.2del | Left.VI | -1.418 | 0.159 | -0.139 | 0.007 | -0.189 |
| 15q11.2del | Left.VIIB | -0.613 | 0.541 | -0.056 | 0.027 | -0.138 |
| 15q11.2del | Left.VIIIA | -2.993 | 0.003 | -0.253 | 0.009 | -0.282 |
| 15q11.2del | Left.VIIIB | -2.158 | 0.033 | -0.187 | 0.026 | -0.134 |
| 15q11.2del | Left.X | -1.457 | 0.148 | -0.154 | -0.004 | -0.099 |
| **15q11.2del** | **Right.Crus.I** | **-3.353** | **0.001** | **-0.320** | **0.001** | **-0.245** |
| 15q11.2del | Right.Crus.II | -2.630 | 0.010 | -0.234 | 0.030 | -0.280 |
| 15q11.2del | Right.I.III | 1.840 | 0.068 | 0.159 | -0.013 | 0.143 |
| 15q11.2del | Right.IX | -0.090 | 0.929 | -0.008 | 0.039 | 0.066 |
| 15q11.2del | Right.V | 0.135 | 0.893 | 0.013 | 0.006 | 0.043 |
| 15q11.2del | Right.VI | -1.164 | 0.246 | -0.109 | 0.022 | -0.059 |
| 15q11.2del | Right.VIIB | -0.992 | 0.323 | -0.096 | 0.014 | -0.086 |
| 15q11.2del | Right.VIIIA | -0.613 | 0.541 | -0.057 | 0.004 | -0.107 |
| 15q11.2del | Right.VIIIB | -2.857 | 0.005 | -0.270 | 0.021 | -0.173 |
| 15q11.2del | Right.X | -1.640 | 0.104 | -0.158 | -0.021 | 0.010 |
| 15q11.2del | Rigt.IV | -2.024 | 0.045 | -0.186 | 0.000 | -0.141 |
| 15q11.2del | Vermis.IX | -1.906 | 0.059 | -0.175 | 0.011 | -0.213 |
| 15q11.2del | Vermis.VI | -1.592 | 0.114 | -0.148 | -0.002 | -0.255 |
| **15q11.2del** | **Vermis.VII** | **-3.233** | **0.002** | **-0.303** | **0.024** | **-0.295** |
| 15q11.2del | Vermis.VIII | -1.372 | 0.173 | -0.113 | 0.025 | -0.012 |
| 15q11.2del | Vermis.X | 0.621 | 0.535 | 0.067 | 0.017 | 0.164 |
| **15q11.2del** | **Total Cerebellar V** | **-2.728** | **0.007** | **-0.273** | **0.050** | **-0.146** |

**Supplementary Table 4**. **Percentage of Extreme Deviations in ROI of CNVs**

| **ROI** | **Pathogenic_CNVs** | **Extreme Negative** | **Extreme Positive** | **Not Extreme** |
| --- | --- | --- | --- | --- |
| Corpus.Medullare | 15q11.2del | 2.5000 | 0.8333 | 96.6667 |
|  | 15q11.2dup | 3.3557 | 2.0134 | 94.6309 |
|  | HC | 2.7433 | 2.0706 | 95.1860 |
| Left.Crus.I | 15q11.2del | 3.3333 | 0.0000 | 96.6667 |
|  | 15q11.2dup | 3.3557 | 2.0134 | 94.6309 |
|  | HC | 2.7749 | 1.5503 | 95.6748 |
| Left.Crus.II | 15q11.2del | 5.0000 | 1.6667 | 93.3333 |
|  | 15q11.2dup | 5.3691 | 1.3423 | 93.2886 |
|  | HC | 2.3387 | 2.1863 | 95.4751 |
| Left.I.III | 15q11.2del | 0.8333 | 5.0000 | 94.1667 |
|  | 15q11.2dup | 2.0134 | 2.0134 | 95.9732 |
|  | HC | 2.3124 | 2.2283 | 95.4593 |
| Left.IV | 15q11.2del | 2.5000 | 3.3333 | 94.1667 |
|  | 15q11.2dup | 2.0134 | 1.3423 | 96.6443 |
|  | HC | 2.0181 | 1.7658 | 96.2161 |
| Left.IX | 15q11.2del | 1.6667 | 3.3333 | 95.0000 |
|  | 15q11.2dup | 6.0403 | 3.3557 | 90.6040 |
|  | HC | 2.8274 | 2.1652 | 95.0074 |
| Left.V | 15q11.2del | 1.6667 | 1.6667 | 96.6667 |
|  | 15q11.2dup | 1.3423 | 0.0000 | 98.6577 |
|  | HC | 1.8341 | 2.0023 | 96.1635 |
| [Left.VI](http://Left.VI) | 15q11.2del | 5.0000 | 2.5000 | 92.5000 |
|  | 15q11.2dup | 6.7114 | 1.3423 | 91.9463 |
|  | HC | 2.6119 | 2.5752 | 94.8129 |
| Left.VIIB | 15q11.2del | 3.3333 | 0.8333 | 95.8333 |
|  | 15q11.2dup | 5.3691 | 0.0000 | 94.6309 |
|  | HC | 2.7959 | 1.9550 | 95.2491 |
| Left.VIIIA | 15q11.2del | 3.3333 | 0.8333 | 95.8333 |
|  | 15q11.2dup | 2.6846 | 1.3423 | 95.9732 |
|  | HC | 2.2756 | 1.9760 | 95.7484 |
| Left.VIIIB | 15q11.2del | 3.3333 | 0.8333 | 95.8333 |
|  | 15q11.2dup | 5.3691 | 2.0134 | 92.6174 |
|  | HC | 2.8432 | 1.5451 | 95.6117 |
| Left.X | 15q11.2del | 6.6667 | 4.1667 | 89.1667 |
|  | 15q11.2dup | 3.3557 | 3.3557 | 93.2886 |
|  | HC | 2.5331 | 1.9708 | 95.4961 |
| Right.Crus.I | 15q11.2del | 6.6667 | 0.0000 | 93.3333 |
|  | 15q11.2dup | 6.7114 | 1.3423 | 91.9463 |
|  | HC | 3.4108 | 2.0706 | 94.5186 |
| Right.Crus.II | 15q11.2del | 2.5000 | 0.0000 | 97.5000 |
|  | 15q11.2dup | 4.6980 | 1.3423 | 93.9597 |
|  | HC | 2.4595 | 1.6029 | 95.9376 |
| Right.I.III | 15q11.2del | 0.8333 | 2.5000 | 96.6667 |
|  | 15q11.2dup | 1.3423 | 2.0134 | 96.6443 |
|  | HC | 1.9392 | 2.3124 | 95.7484 |
| Right.IX | 15q11.2del | 4.1667 | 2.5000 | 93.3333 |
|  | 15q11.2dup | 4.0268 | 2.0134 | 93.9597 |
|  | HC | 2.7749 | 1.8341 | 95.3910 |
| Right.V | 15q11.2del | 3.3333 | 3.3333 | 93.3333 |
|  | 15q11.2dup | 4.0268 | 0.0000 | 95.9732 |
|  | HC | 2.1495 | 2.3860 | 95.4646 |
| [Right.VI](http://Right.VI) | 15q11.2del | 4.1667 | 0.8333 | 95.0000 |
|  | 15q11.2dup | 4.6980 | 2.0134 | 93.2886 |
|  | HC | 2.7118 | 2.5699 | 94.7183 |
| Right.VIIB | 15q11.2del | 2.5000 | 2.5000 | 95.0000 |
|  | 15q11.2dup | 4.0268 | 1.3423 | 94.6309 |
|  | HC | 1.8131 | 1.3244 | 96.8625 |
| Right.VIIIA | 15q11.2del | 3.3333 | 1.6667 | 95.0000 |
|  | 15q11.2dup | 0.6711 | 2.6846 | 96.6443 |
|  | HC | 2.4227 | 2.1442 | 95.4330 |
| Right.VIIIB | 15q11.2del | 4.1667 | 1.6667 | 94.1667 |
|  | 15q11.2dup | 5.3691 | 1.3423 | 93.2886 |
|  | HC | 2.4175 | 1.5766 | 96.0059 |
| Right.X | 15q11.2del | 5.8333 | 3.3333 | 90.8333 |
|  | 15q11.2dup | 4.6980 | 2.6846 | 92.6174 |
|  | HC | 2.5909 | 2.2441 | 95.1650 |
| Rigt.IV | 15q11.2del | 3.3333 | 1.6667 | 95.0000 |
|  | 15q11.2dup | 4.6980 | 0.0000 | 95.3020 |
|  | HC | 1.8236 | 1.7868 | 96.3895 |
| Vermis.IX | 15q11.2del | 4.1667 | 0.8333 | 95.0000 |
|  | 15q11.2dup | 2.0134 | 1.3423 | 96.6443 |
|  | HC | 2.3071 | 1.8236 | 95.8692 |
| [Vermis.VI](http://Vermis.VI) | 15q11.2del | 2.5000 | 2.5000 | 95.0000 |
|  | 15q11.2dup | 3.3557 | 3.3557 | 93.2886 |
|  | HC | 2.5331 | 2.5489 | 94.9180 |
| Vermis.VII | 15q11.2del | 2.5000 | 2.5000 | 95.0000 |
|  | 15q11.2dup | 4.0268 | 4.0268 | 91.9463 |
|  | HC | 2.5121 | 1.9708 | 95.5171 |
| Vermis.VIII | 15q11.2del | 2.5000 | 0.8333 | 96.6667 |
|  | 15q11.2dup | 4.6980 | 1.3423 | 93.9597 |
|  | HC | 2.8484 | 2.2283 | 94.9233 |
| Vermis.X | 15q11.2del | 8.3333 | 5.8333 | 85.8333 |
|  | 15q11.2dup | 2.0134 | 4.0268 | 93.9597 |
|  | HC | 2.9535 | 2.4806 | 94.5659 |
| Total_Cerebel_Vol | 15q11.2del | 5.0000 | 1.6667 | 93.3333 |
|  | 15q11.2dup | 6.7114 | 0.6711 | 92.6174 |
|  | HC | 3.7208 | 1.5924 | 94.6868 |

**Supplementary Table 5. Chi-squared**

The p-values that remain significant after adjustments for multiple comparisons are highlighted in bold.

| **ROI** | **chi stat** | **pvalue** | **dof** |
| --- | --- | --- | --- |
| Corpus.Medullare | 1.1479 | 0.8866 | 4 |
| Left.Crus.I | 2.4101 | 0.6608 | 4 |
| **Left.Crus.II** | **9.9551** | **0.0412** | **4** |
| Left.I.III | 5.3310 | 0.2550 | 4 |
| Left.IV | 2.0020 | 0.7354 | 4 |
| Left.IX | 7.9497 | 0.0934 | 4 |
| Left.V | 3.3595 | 0.4996 | 4 |
| [**Left.VI**](http://Left.VI) | **12.9087** | **0.0117** | **4** |
| Left.VIIB | 7.2748 | 0.1221 | 4 |
| Left.VIIIA | 1.7778 | 0.7766 | 4 |
| Left.VIIIB | 4.1284 | 0.3889 | 4 |
| **Left.X** | **13.1575** | **0.0105** | **4** |
| **Right.Crus.I** | **11.2406** | **0.0240** | **4** |
| Right.Crus.II | 5.0662 | 0.2806 | 4 |
| Right.I.III | 1.1221 | 0.8908 | 4 |
| Right.IX | 2.0414 | 0.7282 | 4 |
| Right.V | 7.2266 | 0.1244 | 4 |
| [Right.VI](http://Right.VI) | 4.6475 | 0.3254 | 4 |
| Right.VIIB | 5.5966 | 0.2314 | 4 |
| Right.VIIIA | 2.6555 | 0.6170 | 4 |
| Right.VIIIB | 6.9173 | 0.1403 | 4 |
| Right.X | 8.3071 | 0.0810 | 4 |
| **Rigt.IV** | **10.7188** | **0.0299** | **4** |
| Vermis.IX | 2.6872 | 0.6115 | 4 |
| [Vermis.VI](http://Vermis.VI) | 0.8143 | 0.9365 | 4 |
| Vermis.VII | 4.8434 | 0.3037 | 4 |
| Vermis.VIII | 3.4387 | 0.4873 | 4 |
| **Vermis.X** | **19.7221** | **0.0006** | **4** |
| Total_Cerebel_Vol | 4.9112 | 0.2965 | 4 |

**Supplementary Table 6. Post-hoc of Z-test**

The p-values that remain significant after adjustments for multiple comparisons are highlighted in bold.

| **ROI** | **Group 1** | **Group 2** | **Category** | **Z-score** | **P-value** |
| --- | --- | --- | --- | --- | --- |
| Corpus.Medullare | 15q11.2del | 15q11.2dup | Extreme Negative | -0.4107 | 0.6813 |
|  | 15q11.2del | 15q11.2dup | Extreme Positive | -0.7949 | 0.4267 |
|  | 15q11.2del | 15q11.2dup | Not Extreme | 0.8040 | 0.4214 |
|  | 15q11.2del | HC | Extreme Negative | -0.1627 | 0.8707 |
|  | 15q11.2del | HC | Extreme Positive | -0.9506 | 0.3418 |
|  | 15q11.2del | HC | Not Extreme | 0.7560 | 0.4496 |
|  | 15q11.2dup | HC | Extreme Negative | 0.4555 | 0.6488 |
|  | 15q11.2dup | HC | Extreme Positive | -0.0489 | 0.9610 |
|  | 15q11.2dup | HC | Not Extreme | -0.3152 | 0.7526 |
| Left.Crus.I | 15q11.2del | 15q11.2dup | Extreme Negative | -0.0101 | 0.9919 |
|  | 15q11.2del | 15q11.2dup | Extreme Positive | -1.5631 | 0.1180 |
|  | 15q11.2del | 15q11.2dup | Not Extreme | 0.8040 | 0.4214 |
|  | 15q11.2del | HC | Extreme Negative | 0.3711 | 0.7106 |
|  | 15q11.2del | HC | Extreme Positive | -1.3746 | 0.1693 |
|  | 15q11.2del | HC | Not Extreme | 0.5328 | 0.5942 |
|  | 15q11.2dup | HC | Extreme Negative | 0.4296 | 0.6675 |
|  | 15q11.2dup | HC | Extreme Positive | 0.4552 | 0.6489 |
|  | 15q11.2dup | HC | Not Extreme | -0.6234 | 0.5330 |
| Left.Crus.II | 15q11.2del | 15q11.2dup | Extreme Negative | -0.1355 | 0.8922 |
|  | 15q11.2del | 15q11.2dup | Extreme Positive | 0.2185 | 0.8270 |
|  | 15q11.2del | 15q11.2dup | Not Extreme | 0.0146 | 0.9884 |
|  | 15q11.2del | HC | Extreme Negative | 1.9164 | 0.0553 |
|  | 15q11.2del | HC | Extreme Positive | -0.3883 | 0.6978 |
|  | 15q11.2del | HC | Not Extreme | -1.1237 | 0.2612 |
|  | 15q11.2dup | HC | Extreme Negative | 2.4263 | 0.0153 |
|  | 15q11.2dup | HC | Extreme Positive | -0.7028 | 0.4822 |
|  | 15q11.2dup | HC | Not Extreme | -1.2768 | 0.2017 |
| Left.I.III | 15q11.2del | 15q11.2dup | Extreme Negative | -0.7949 | 0.4267 |
|  | 15q11.2del | 15q11.2dup | Extreme Positive | 1.3540 | 0.1757 |
|  | 15q11.2del | 15q11.2dup | Not Extreme | -0.6868 | 0.4922 |
|  | 15q11.2del | HC | Extreme Negative | -1.0767 | 0.2816 |
|  | 15q11.2del | HC | Extreme Positive | 2.0428 | 0.0411 |
|  | 15q11.2del | HC | Not Extreme | -0.6774 | 0.4981 |
|  | 15q11.2dup | HC | Extreme Negative | -0.2420 | 0.8088 |
|  | 15q11.2dup | HC | Extreme Positive | -0.1771 | 0.8595 |
|  | 15q11.2dup | HC | Not Extreme | 0.3002 | 0.7640 |
| Left.IV | 15q11.2del | 15q11.2dup | Extreme Negative | 0.2686 | 0.7882 |
|  | 15q11.2del | 15q11.2dup | Extreme Positive | 1.0992 | 0.2717 |
|  | 15q11.2del | 15q11.2dup | Not Extreme | -0.9785 | 0.3279 |
|  | 15q11.2del | HC | Extreme Negative | 0.3740 | 0.7084 |
|  | 15q11.2del | HC | Extreme Positive | 1.2961 | 0.1949 |
|  | 15q11.2del | HC | Not Extreme | -1.1710 | 0.2416 |
|  | 15q11.2dup | HC | Extreme Negative | -0.0040 | 0.9968 |
|  | 15q11.2dup | HC | Extreme Positive | -0.3914 | 0.6955 |
|  | 15q11.2dup | HC | Not Extreme | 0.2730 | 0.7849 |
| Left.IX | 15q11.2del | 15q11.2dup | Extreme Negative | -1.8005 | 0.0718 |
|  | 15q11.2del | 15q11.2dup | Extreme Positive | -0.0101 | 0.9919 |
|  | 15q11.2del | 15q11.2dup | Not Extreme | 1.3662 | 0.1719 |
|  | 15q11.2del | HC | Extreme Negative | -0.7657 | 0.4439 |
|  | 15q11.2del | HC | Extreme Positive | 0.8750 | 0.3816 |
|  | 15q11.2del | HC | Not Extreme | -0.0037 | 0.9971 |
|  | 15q11.2dup | HC | Extreme Negative | 2.3468 | 0.0189 |
|  | 15q11.2dup | HC | Extreme Positive | 0.9925 | 0.3210 |
|  | 15q11.2dup | HC | Not Extreme | -2.4504 | 0.0143 |
| Left.V | 15q11.2del | 15q11.2dup | Extreme Negative | 0.2185 | 0.8270 |
|  | 15q11.2del | 15q11.2dup | Extreme Positive | 1.5818 | 0.1137 |
|  | 15q11.2del | 15q11.2dup | Not Extreme | -1.0992 | 0.2717 |
|  | 15q11.2del | HC | Extreme Negative | -0.1363 | 0.8916 |
|  | 15q11.2del | HC | Extreme Positive | -0.2618 | 0.7935 |
|  | 15q11.2del | HC | Not Extreme | 0.2862 | 0.7748 |
|  | 15q11.2dup | HC | Extreme Negative | -0.4462 | 0.6555 |
|  | 15q11.2dup | HC | Extreme Positive | -1.7447 | 0.0810 |
|  | 15q11.2dup | HC | Not Extreme | 1.5827 | 0.1135 |
| [Left.VI](http://Left.VI) | 15q11.2del | 15q11.2dup | Extreme Negative | -0.5899 | 0.5552 |
|  | 15q11.2del | 15q11.2dup | Extreme Positive | 0.6988 | 0.4847 |
|  | 15q11.2del | 15q11.2dup | Not Extreme | 0.1683 | 0.8664 |
|  | 15q11.2del | HC | Extreme Negative | 1.6305 | 0.1030 |
|  | 15q11.2del | HC | Extreme Positive | -0.0518 | 0.9587 |
|  | 15q11.2del | HC | Not Extreme | -1.1374 | 0.2554 |
|  | **15q11.2dup** | **HC** | **Extreme Negative** | **3.1069** | **0.0019** |
|  | 15q11.2dup | HC | Extreme Positive | -0.9481 | 0.3431 |
|  | 15q11.2dup | HC | Not Extreme | -1.5685 | 0.1168 |
| Left.VIIB | 15q11.2del | 15q11.2dup | Extreme Negative | -0.8040 | 0.4214 |
|  | 15q11.2del | 15q11.2dup | Extreme Positive | 1.1164 | 0.2643 |
|  | 15q11.2del | 15q11.2dup | Not Extreme | 0.4571 | 0.6476 |
|  | 15q11.2del | HC | Extreme Negative | 0.3558 | 0.7220 |
|  | 15q11.2del | HC | Extreme Positive | -0.8863 | 0.3755 |
|  | 15q11.2del | HC | Not Extreme | 0.3000 | 0.7642 |
|  | 15q11.2dup | HC | Extreme Negative | 1.8914 | 0.0586 |
|  | 15q11.2dup | HC | Extreme Positive | -1.7235 | 0.0848 |
|  | 15q11.2dup | HC | Not Extreme | -0.3532 | 0.7239 |
| Left.VIIIA | 15q11.2del | 15q11.2dup | Extreme Negative | 0.3114 | 0.7555 |
|  | 15q11.2del | 15q11.2dup | Extreme Positive | -0.3951 | 0.6928 |
|  | 15q11.2del | 15q11.2dup | Not Extreme | -0.0576 | 0.9541 |
|  | 15q11.2del | HC | Extreme Negative | 0.7735 | 0.4392 |
|  | 15q11.2del | HC | Extreme Positive | -0.8982 | 0.3691 |
|  | 15q11.2del | HC | Not Extreme | 0.0460 | 0.9633 |
|  | 15q11.2dup | HC | Extreme Negative | 0.3332 | 0.7390 |
|  | 15q11.2dup | HC | Extreme Positive | -0.5544 | 0.5793 |
|  | 15q11.2dup | HC | Not Extreme | 0.1355 | 0.8922 |
| Left.VIIIB | 15q11.2del | 15q11.2dup | Extreme Negative | -0.8040 | 0.4214 |
|  | 15q11.2del | 15q11.2dup | Extreme Positive | -0.7949 | 0.4267 |
|  | 15q11.2del | 15q11.2dup | Not Extreme | 1.1085 | 0.2676 |
|  | 15q11.2del | HC | Extreme Negative | 0.3219 | 0.7475 |
|  | 15q11.2del | HC | Extreme Positive | -0.6311 | 0.5280 |
|  | 15q11.2del | HC | Not Extreme | 0.1182 | 0.9059 |
|  | 15q11.2dup | HC | Extreme Negative | 1.8418 | 0.0655 |
|  | 15q11.2dup | HC | Extreme Positive | 0.4612 | 0.6447 |
|  | 15q11.2dup | HC | Not Extreme | -1.7729 | 0.0762 |
| Left.X | 15q11.2del | 15q11.2dup | Extreme Negative | 1.2587 | 0.2081 |
|  | 15q11.2del | 15q11.2dup | Extreme Positive | 0.3495 | 0.7267 |
|  | 15q11.2del | 15q11.2dup | Not Extreme | -1.2018 | 0.2294 |
|  | 15q11.2del | HC | Extreme Negative | 2.8585 | 0.0043 |
|  | 15q11.2del | HC | Extreme Positive | 1.7193 | 0.0856 |
|  | 15q11.2del | HC | Not Extreme | -3.3189 | 0.0009 |
|  | 15q11.2dup | HC | Extreme Negative | 0.6358 | 0.5249 |
|  | 15q11.2dup | HC | Extreme Positive | 1.2083 | 0.2269 |
|  | 15q11.2dup | HC | Not Extreme | -1.2919 | 0.1964 |
| Right.Crus.I | 15q11.2del | 15q11.2dup | Extreme Negative | -0.0146 | 0.9884 |
|  | 15q11.2del | 15q11.2dup | Extreme Positive | -1.2739 | 0.2027 |
|  | 15q11.2del | 15q11.2dup | Not Extreme | 0.4311 | 0.6664 |
|  | 15q11.2del | HC | Extreme Negative | 1.9532 | 0.0508 |
|  | 15q11.2del | HC | Extreme Positive | -1.5928 | 0.1112 |
|  | 15q11.2del | HC | Not Extreme | -0.5683 | 0.5698 |
|  | 15q11.2dup | HC | Extreme Negative | 2.2031 | 0.0276 |
|  | 15q11.2dup | HC | Extreme Positive | -0.6227 | 0.5334 |
|  | 15q11.2dup | HC | Not Extreme | -1.3717 | 0.1701 |
| Right.Crus.II | 15q11.2del | 15q11.2dup | Extreme Negative | -0.9472 | 0.3435 |
|  | 15q11.2del | 15q11.2dup | Extreme Positive | -1.2739 | 0.2027 |
|  | 15q11.2del | 15q11.2dup | Not Extreme | 1.3981 | 0.1621 |
|  | 15q11.2del | HC | Extreme Negative | 0.0285 | 0.9772 |
|  | 15q11.2del | HC | Extreme Positive | -1.3981 | 0.1621 |
|  | 15q11.2del | HC | Not Extreme | 0.8652 | 0.3869 |
|  | 15q11.2dup | HC | Extreme Negative | 1.7512 | 0.0799 |
|  | 15q11.2dup | HC | Extreme Positive | -0.2525 | 0.8007 |
|  | 15q11.2dup | HC | Not Extreme | -1.2160 | 0.2240 |
| Right.I.III | 15q11.2del | 15q11.2dup | Extreme Negative | -0.3951 | 0.6928 |
|  | 15q11.2del | 15q11.2dup | Extreme Positive | 0.2686 | 0.7882 |
|  | 15q11.2del | 15q11.2dup | Not Extreme | 0.0101 | 0.9919 |
|  | 15q11.2del | HC | Extreme Negative | -0.8773 | 0.3803 |
|  | 15q11.2del | HC | Extreme Positive | 0.1363 | 0.8916 |
|  | 15q11.2del | HC | Not Extreme | 0.4973 | 0.6190 |
|  | 15q11.2dup | HC | Extreme Negative | -0.5270 | 0.5982 |
|  | 15q11.2dup | HC | Extreme Positive | -0.2420 | 0.8088 |
|  | 15q11.2dup | HC | Not Extreme | 0.5403 | 0.5890 |
| Right.IX | 15q11.2del | 15q11.2dup | Extreme Negative | 0.0576 | 0.9541 |
|  | 15q11.2del | 15q11.2dup | Extreme Positive | 0.2686 | 0.7882 |
|  | 15q11.2del | 15q11.2dup | Not Extreme | -0.2099 | 0.8338 |
|  | 15q11.2del | HC | Extreme Negative | 0.9239 | 0.3555 |
|  | 15q11.2del | HC | Extreme Positive | 0.5413 | 0.5883 |
|  | 15q11.2del | HC | Not Extreme | -1.0702 | 0.2845 |
|  | 15q11.2dup | HC | Extreme Negative | 0.9252 | 0.3548 |
|  | 15q11.2dup | HC | Extreme Positive | 0.1624 | 0.8710 |
|  | 15q11.2dup | HC | Not Extreme | -0.8290 | 0.4071 |
| Right.V | 15q11.2del | 15q11.2dup | Extreme Negative | -0.2989 | 0.7650 |
|  | 15q11.2del | 15q11.2dup | Extreme Positive | 2.2454 | 0.0247 |
|  | 15q11.2del | 15q11.2dup | Not Extreme | -0.9689 | 0.3326 |
|  | 15q11.2del | HC | Extreme Negative | 0.8899 | 0.3735 |
|  | 15q11.2del | HC | Extreme Positive | 0.6771 | 0.4984 |
|  | 15q11.2del | HC | Not Extreme | -1.1169 | 0.2640 |
|  | 15q11.2dup | HC | Extreme Negative | 1.5688 | 0.1167 |
|  | 15q11.2dup | HC | Extreme Positive | -1.9082 | 0.0564 |
|  | 15q11.2dup | HC | Not Extreme | 0.2973 | 0.7662 |
| [Right.VI](http://Right.VI) | 15q11.2del | 15q11.2dup | Extreme Negative | -0.2098 | 0.8338 |
|  | 15q11.2del | 15q11.2dup | Extreme Positive | -0.7949 | 0.4267 |
|  | 15q11.2del | 15q11.2dup | Not Extreme | 0.5899 | 0.5552 |
|  | 15q11.2del | HC | Extreme Negative | 0.9765 | 0.3288 |
|  | 15q11.2del | HC | Extreme Positive | -1.2009 | 0.2298 |
|  | 15q11.2del | HC | Not Extreme | 0.1376 | 0.8906 |
|  | 15q11.2dup | HC | Extreme Negative | 1.4827 | 0.1381 |
|  | 15q11.2dup | HC | Extreme Positive | -0.4280 | 0.6687 |
|  | 15q11.2dup | HC | Not Extreme | -0.7765 | 0.4375 |
| Right.VIIB | 15q11.2del | 15q11.2dup | Extreme Negative | -0.6922 | 0.4888 |
|  | 15q11.2del | 15q11.2dup | Extreme Positive | 0.6988 | 0.4847 |
|  | 15q11.2del | 15q11.2dup | Not Extreme | 0.1355 | 0.8922 |
|  | 15q11.2del | HC | Extreme Negative | 0.5615 | 0.5744 |
|  | 15q11.2del | HC | Extreme Positive | 1.1200 | 0.2627 |
|  | 15q11.2del | HC | Not Extreme | -1.1646 | 0.2442 |
|  | 15q11.2dup | HC | Extreme Negative | 2.0080 | 0.0446 |
|  | 15q11.2dup | HC | Extreme Positive | 0.0191 | 0.9848 |
|  | 15q11.2dup | HC | Not Extreme | -1.5524 | 0.1206 |
| Right.VIIIA | 15q11.2del | 15q11.2dup | Extreme Negative | 1.6070 | 0.1081 |
|  | 15q11.2del | 15q11.2dup | Extreme Positive | -0.5620 | 0.5741 |
|  | 15q11.2del | 15q11.2dup | Not Extreme | -0.6769 | 0.4985 |
|  | 15q11.2del | HC | Extreme Negative | 0.6460 | 0.5183 |
|  | 15q11.2del | HC | Extreme Positive | -0.3603 | 0.7187 |
|  | 15q11.2del | HC | Not Extreme | -0.2265 | 0.8209 |
|  | 15q11.2dup | HC | Extreme Negative | -1.3890 | 0.1648 |
|  | 15q11.2dup | HC | Extreme Positive | 0.4531 | 0.6504 |
|  | 15q11.2dup | HC | Not Extreme | 0.7061 | 0.4801 |
| Right.VIIIB | 15q11.2del | 15q11.2dup | Extreme Negative | -0.4571 | 0.6476 |
|  | 15q11.2del | 15q11.2dup | Extreme Positive | 0.2185 | 0.8270 |
|  | 15q11.2del | 15q11.2dup | Not Extreme | 0.2942 | 0.7686 |
|  | 15q11.2del | HC | Extreme Negative | 1.2409 | 0.2146 |
|  | 15q11.2del | HC | Extreme Positive | 0.0789 | 0.9371 |
|  | 15q11.2del | HC | Not Extreme | -1.0242 | 0.3057 |
|  | 15q11.2dup | HC | Extreme Negative | 2.3259 | 0.0200 |
|  | 15q11.2dup | HC | Extreme Positive | -0.2289 | 0.8190 |
|  | 15q11.2dup | HC | Not Extreme | -1.6830 | 0.0924 |
| Right.X | 15q11.2del | 15q11.2dup | Extreme Negative | 0.4167 | 0.6769 |
|  | 15q11.2del | 15q11.2dup | Extreme Positive | 0.3114 | 0.7555 |
|  | 15q11.2del | 15q11.2dup | Not Extreme | -0.5308 | 0.5956 |
|  | 15q11.2del | HC | Extreme Negative | 2.2203 | 0.0264 |
|  | 15q11.2del | HC | Extreme Positive | 0.8019 | 0.4226 |
|  | 15q11.2del | HC | Not Extreme | -2.1993 | 0.0279 |
|  | 15q11.2dup | HC | Extreme Negative | 1.6078 | 0.1079 |
|  | 15q11.2dup | HC | Extreme Positive | 0.3614 | 0.7178 |
|  | 15q11.2dup | HC | Not Extreme | -1.4413 | 0.1495 |
| Rigt.IV | 15q11.2del | 15q11.2dup | Extreme Negative | -0.5618 | 0.5743 |
|  | 15q11.2del | 15q11.2dup | Extreme Positive | 1.5818 | 0.1137 |
|  | 15q11.2del | 15q11.2dup | Not Extreme | -0.1148 | 0.9086 |
|  | 15q11.2del | HC | Extreme Negative | 1.2290 | 0.2191 |
|  | 15q11.2del | HC | Extreme Positive | -0.0991 | 0.9211 |
|  | 15q11.2del | HC | Not Extreme | -0.8124 | 0.4165 |
|  | 15q11.2dup | HC | Extreme Negative | 2.5964 | 0.0094 |
|  | 15q11.2dup | HC | Extreme Positive | -1.6463 | 0.0997 |
|  | 15q11.2dup | HC | Not Extreme | -0.7080 | 0.4789 |
| Vermis.IX | 15q11.2del | 15q11.2dup | Extreme Negative | 1.0334 | 0.3014 |
|  | 15q11.2del | 15q11.2dup | Extreme Positive | -0.3951 | 0.6928 |
|  | 15q11.2del | 15q11.2dup | Not Extreme | -0.6769 | 0.4985 |
|  | 15q11.2del | HC | Extreme Negative | 1.3493 | 0.1773 |
|  | 15q11.2del | HC | Extreme Positive | -0.8096 | 0.4182 |
|  | 15q11.2del | HC | Not Extreme | -0.4767 | 0.6336 |
|  | 15q11.2dup | HC | Extreme Negative | -0.2380 | 0.8119 |
|  | 15q11.2dup | HC | Extreme Positive | -0.4378 | 0.6615 |
|  | 15q11.2dup | HC | Not Extreme | 0.4739 | 0.6356 |
| [Vermis.VI](http://Vermis.VI) | 15q11.2del | 15q11.2dup | Extreme Negative | -0.4107 | 0.6813 |
|  | 15q11.2del | 15q11.2dup | Extreme Positive | -0.4107 | 0.6813 |
|  | 15q11.2del | 15q11.2dup | Not Extreme | 0.5899 | 0.5552 |
|  | 15q11.2del | HC | Extreme Negative | -0.0230 | 0.9816 |
|  | 15q11.2del | HC | Extreme Positive | -0.0339 | 0.9730 |
|  | 15q11.2del | HC | Not Extreme | 0.0408 | 0.9675 |
|  | 15q11.2dup | HC | Extreme Negative | 0.6358 | 0.5249 |
|  | 15q11.2dup | HC | Extreme Positive | 0.6217 | 0.5341 |
|  | 15q11.2dup | HC | Not Extreme | -0.9010 | 0.3676 |
| Vermis.VII | 15q11.2del | 15q11.2dup | Extreme Negative | -0.6922 | 0.4888 |
|  | 15q11.2del | 15q11.2dup | Extreme Positive | -0.6922 | 0.4888 |
|  | 15q11.2del | 15q11.2dup | Not Extreme | 0.9963 | 0.3191 |
|  | 15q11.2del | HC | Extreme Negative | -0.0084 | 0.9933 |
|  | 15q11.2del | HC | Extreme Positive | 0.4154 | 0.6778 |
|  | 15q11.2del | HC | Not Extreme | -0.2728 | 0.7850 |
|  | 15q11.2dup | HC | Extreme Negative | 1.1743 | 0.2403 |
|  | 15q11.2dup | HC | Extreme Positive | 1.7915 | 0.0732 |
|  | 15q11.2dup | HC | Not Extreme | -2.0921 | 0.0364 |
| Vermis.VIII | 15q11.2del | 15q11.2dup | Extreme Negative | -0.9472 | 0.3435 |
|  | 15q11.2del | 15q11.2dup | Extreme Positive | -0.3951 | 0.6928 |
|  | 15q11.2del | 15q11.2dup | Not Extreme | 1.0291 | 0.3034 |
|  | 15q11.2del | HC | Extreme Negative | -0.2288 | 0.8190 |
|  | 15q11.2del | HC | Extreme Positive | -1.0340 | 0.3011 |
|  | 15q11.2del | HC | Not Extreme | 0.8681 | 0.3853 |
|  | 15q11.2dup | HC | Extreme Negative | 1.3486 | 0.1775 |
|  | 15q11.2dup | HC | Extreme Positive | -0.7310 | 0.4648 |
|  | 15q11.2dup | HC | Not Extreme | -0.5333 | 0.5938 |
| Vermis.X | 15q11.2del | 15q11.2dup | Extreme Negative | 2.4026 | 0.0163 |
|  | 15q11.2del | 15q11.2dup | Extreme Positive | 0.6868 | 0.4922 |
|  | 15q11.2del | 15q11.2dup | Not Extreme | -2.2422 | 0.0250 |
|  | **15q11.2del** | **HC** | **Extreme Negative** | **3.4510** | **0.0006** |
|  | 15q11.2del | HC | Extreme Positive | 2.3444 | 0.0191 |
|  | 15q11.2del | HC | Not Extreme | -4.1868 | 0.0000 |
|  | 15q11.2dup | HC | Extreme Negative | -0.6760 | 0.4990 |
|  | 15q11.2dup | HC | Extreme Positive | 1.2060 | 0.2278 |
|  | 15q11.2dup | HC | Not Extreme | -0.3250 | 0.7452 |
| Total_Cerebel_Vol | 15q11.2del | 15q11.2dup | Extreme Negative | -0.5899 | 0.5552 |
|  | 15q11.2del | 15q11.2dup | Extreme Positive | 0.7729 | 0.4396 |
|  | 15q11.2del | 15q11.2dup | Not Extreme | 0.2278 | 0.8198 |
|  | 15q11.2del | HC | Extreme Negative | 0.7373 | 0.4610 |
|  | 15q11.2del | HC | Extreme Positive | 0.0648 | 0.9483 |
|  | 15q11.2del | HC | Not Extreme | -0.6584 | 0.5103 |
|  | 15q11.2dup | HC | Extreme Negative | 1.9154 | 0.0554 |
|  | 15q11.2dup | HC | Extreme Positive | -0.8968 | 0.3698 |
|  | 15q11.2dup | HC | Not Extreme | -1.1202 | 0.2626 |

**Supplementary Table 7**. **Dose effect of CNVs**

The p-values that remain significant after adjustments for multiple comparisons are highlighted in bold.

| **ROI** | **param** | **P-value** | **T-stat Del vs Non-carrier** | **P-value Del vs Non-carrier** | **T-stat Dup vs Non-carrier** | **P-value Dup vs Non-carrier** | **T-stat Dup vs Del** | **P-value Dup vs Del** |
| --- | --- | --- | --- | --- | --- | --- | --- | --- |
| Corpus.Medullare | -0.206 | 0.095 | -2.642 | 0.009 | -0.087 | 0.931 | 1.878 | 0.061 |
| Left.Crus.I | -0.043 | 0.849 | -2.391 | 0.018 | -1.827 | 0.070 | 0.496 | 0.620 |
| Left.Crus.II | -0.064 | 0.587 | -1.716 | 0.089 | -0.725 | 0.470 | 0.722 | 0.471 |
| Left.I.III | 0.303 | 0.011 | 2.178 | 0.031 | -1.480 | 0.141 | -2.609 | 0.010 |
| Left.IV | -0.017 | 0.949 | -0.579 | 0.564 | -0.444 | 0.658 | 0.137 | 0.891 |
| Left.IX | 0.085 | 0.602 | -1.130 | 0.260 | -1.487 | 0.139 | -0.345 | 0.731 |
| Left.V | 0.022 | 0.923 | -1.729 | 0.086 | -1.738 | 0.084 | 0.151 | 0.880 |
| Left.VI | 0.165 | 0.217 | -1.418 | 0.159 | -3.121 | 0.002 | -0.927 | 0.355 |
| Left.VIIB | 0.150 | 0.217 | -0.613 | 0.541 | -2.277 | 0.024 | -1.033 | 0.303 |
| Left.VIIIA | -0.162 | 0.146 | -2.993 | 0.003 | -0.712 | 0.478 | 1.737 | 0.084 |
| Left.VIIIB | -0.165 | 0.198 | -2.158 | 0.033 | -0.069 | 0.945 | 1.461 | 0.145 |
| Left.X | -0.150 | 0.310 | -1.457 | 0.148 | 0.002 | 0.998 | 1.117 | 0.265 |
| Right.Crus.I | -0.157 | 0.339 | -3.353 | 0.001 | -1.627 | 0.106 | 1.336 | 0.183 |
| Right.Crus.II | -0.053 | 0.728 | -2.630 | 0.010 | -1.518 | 0.131 | 0.650 | 0.516 |
| Right.I.III | 0.371 | 0.001 | 1.840 | 0.068 | -2.764 | 0.006 | -3.228 | 0.001 |
| Right.IX | 0.111 | 0.441 | -0.090 | 0.929 | -1.049 | 0.296 | -0.695 | 0.488 |
| Right.V | 0.144 | 0.255 | 0.135 | 0.893 | -1.464 | 0.145 | -1.041 | 0.299 |
| Right.VI | 0.108 | 0.411 | -1.164 | 0.246 | -2.266 | 0.025 | -0.583 | 0.561 |
| Right.VIIB | 0.096 | 0.442 | -0.992 | 0.323 | -1.957 | 0.052 | -0.564 | 0.573 |
| Right.VIIIA | -0.105 | 0.347 | -0.613 | 0.541 | 0.712 | 0.477 | 0.929 | 0.354 |
| Right.VIIIB | -0.187 | 0.143 | -2.857 | 0.005 | -0.535 | 0.594 | 1.727 | 0.085 |
| Right.X | -0.022 | 0.981 | -1.640 | 0.104 | -1.478 | 0.141 | 0.199 | 0.843 |
| Rigt.IV | 0.148 | 0.183 | -2.024 | 0.045 | -3.786 | 0.000 | -0.951 | 0.343 |
| Vermis.IX | -0.190 | 0.125 | -1.906 | 0.059 | 0.337 | 0.737 | 1.648 | 0.101 |
| Vermis.VI | -0.151 | 0.262 | -1.592 | 0.114 | 0.156 | 0.876 | 1.233 | 0.219 |
| Vermis.VII | -0.271 | 0.030 | -3.233 | 0.002 | 0.109 | 0.913 | 2.425 | 0.016 |
| Vermis.VIII | -0.131 | 0.258 | -1.372 | 0.173 | 0.346 | 0.730 | 1.212 | 0.227 |
| Vermis.X | -0.064 | 0.627 | 0.621 | 0.535 | 1.391 | 0.166 | 0.341 | 0.733 |
| Total_Cerebel_Vol | -0.047 | 0.756 | -2.692 | 0.008 | -1.909 | 0.058 | 0.647 | 0.518 |

**Supplementary Table 8**. **Percentage of Extreme Deviations in Subcortical of CNVs**

| **cnv** | **roi** | **Percentage of positive outliers** | **Percentage of negative outliers** |
| --- | --- | --- | --- |
| 15q11.2del | lh_MeanThickness_thickness | 4.167 | 1.667 |
| 15q11.2del | rh_MeanThickness_thickness | 5.000 | 1.667 |
| 15q11.2del | Left-Cerebellum-White-Matter | 3.333 | 2.500 |
| 15q11.2del | Left-Cerebellum-Cortex | 2.500 | 2.500 |
| 15q11.2del | Left-Thalamus-Proper | 2.500 | 0.833 |
| 15q11.2del | Left-Caudate | 0.833 | 0.833 |
| 15q11.2del | Left-Putamen | 1.667 | 0.833 |
| 15q11.2del | Left-Pallidum | 1.667 | 0.000 |
| 15q11.2del | Left-Hippocampus | 3.333 | 1.667 |
| 15q11.2del | Left-Amygdala | 3.333 | 1.667 |
| 15q11.2del | Left-Accumbens-area | 0.833 | 0.833 |
| 15q11.2del | Right-Cerebellum-White-Matter | 2.500 | 2.500 |
| 15q11.2del | Right-Cerebellum-Cortex | 3.333 | 2.500 |
| 15q11.2del | Right-Thalamus-Proper | 4.167 | 1.667 |
| 15q11.2del | Right-Caudate | 1.667 | 0.833 |
| 15q11.2del | Right-Putamen | 1.667 | 0.000 |
| 15q11.2del | Right-Pallidum | 1.667 | 1.667 |
| 15q11.2del | Right-Hippocampus | 4.167 | 1.667 |
| 15q11.2del | Right-Amygdala | 5.833 | 0.000 |
| 15q11.2del | Right-Accumbens-area | 0.000 | 0.833 |
| 15q11.2dup | lh_MeanThickness_thickness | 1.351 | 1.351 |
| 15q11.2dup | rh_MeanThickness_thickness | 1.351 | 2.703 |
| 15q11.2dup | Left-Cerebellum-White-Matter | 2.703 | 1.351 |
| 15q11.2dup | Left-Cerebellum-Cortex | 1.351 | 3.378 |
| 15q11.2dup | Left-Thalamus-Proper | 2.027 | 0.676 |
| 15q11.2dup | Left-Caudate | 2.027 | 1.351 |
| 15q11.2dup | Left-Putamen | 2.027 | 2.027 |
| 15q11.2dup | Left-Pallidum | 2.027 | 0.000 |
| 15q11.2dup | Left-Hippocampus | 3.378 | 3.378 |
| 15q11.2dup | Left-Amygdala | 2.703 | 1.351 |
| 15q11.2dup | Left-Accumbens-area | 4.054 | 0.000 |
| 15q11.2dup | Right-Cerebellum-White-Matter | 2.027 | 2.027 |
| 15q11.2dup | Right-Cerebellum-Cortex | 1.351 | 2.703 |
| 15q11.2dup | Right-Thalamus-Proper | 3.378 | 0.676 |
| 15q11.2dup | Right-Caudate | 2.703 | 0.000 |
| 15q11.2dup | Right-Putamen | 2.703 | 1.351 |
| 15q11.2dup | Right-Pallidum | 2.027 | 2.027 |
| 15q11.2dup | Right-Hippocampus | 2.027 | 2.027 |
| 15q11.2dup | Right-Amygdala | 7.432 | 1.351 |
| 15q11.2dup | Right-Accumbens-area | 4.054 | 0.676 |

**Supplementary Table 9**. **Percentage of Extreme Deviations in Surface Area of CNVs**

a) **Right hemisphere for 15q11.2**

| **cnv** | **roi** | **Percentage of positive outliers** | **ercentage of negative outliers** |
| --- | --- | --- | --- |
| 15q11.2del | G_and_S_frontomargin | 2.439 | 3.252 |
| 15q11.2del | G_and_S_occipital_inf | 4.065 | 1.626 |
| 15q11.2del | G_and_S_paracentral | 3.252 | 0.813 |
| 15q11.2del | G_and_S_subcentral | 0.813 | 2.439 |
| 15q11.2del | G_and_S_transv_frontopol | 7.317 | 1.626 |
| 15q11.2del | G_and_S_cingul-Ant | 1.626 | 5.691 |
| 15q11.2del | G_and_S_cingul-Mid-Ant | 0.813 | 2.439 |
| 15q11.2del | G_and_S_cingul-Mid-Post | 2.439 | 3.252 |
| 15q11.2del | G_cingul-Post-dorsal | 2.439 | 4.878 |
| 15q11.2del | G_cingul-Post-ventral | 5.691 | 3.252 |
| 15q11.2del | G_cuneus | 1.626 | 1.626 |
| 15q11.2del | G_front_inf-Opercular | 2.439 | 0.000 |
| 15q11.2del | G_front_inf-Orbital | 1.626 | 2.439 |
| 15q11.2del | G_front_inf-Triangul | 0.000 | 3.252 |
| 15q11.2del | G_front_middle | 0.000 | 3.252 |
| 15q11.2del | G_front_sup | 2.439 | 0.813 |
| 15q11.2del | G_Ins_lg_and_S_cent_ins | 3.252 | 0.813 |
| 15q11.2del | G_insular_short | 2.439 | 0.813 |
| 15q11.2del | G_occipital_middle | 0.813 | 4.065 |
| 15q11.2del | G_occipital_sup | 3.252 | 3.252 |
| 15q11.2del | G_oc-temp_lat-fusifor | 1.626 | 2.439 |
| 15q11.2del | G_oc-temp_med-Lingual | 3.252 | 3.252 |
| 15q11.2del | G_oc-temp_med-Parahip | 3.252 | 1.626 |
| 15q11.2del | G_orbital | 0.813 | 4.065 |
| 15q11.2del | G_pariet_inf-Angular | 0.813 | 3.252 |
| 15q11.2del | G_pariet_inf-Supramar | 3.252 | 3.252 |
| 15q11.2del | G_parietal_sup | 0.813 | 5.691 |
| 15q11.2del | G_postcentral | 1.626 | 3.252 |
| 15q11.2del | G_precentral | 2.439 | 0.813 |
| 15q11.2del | G_precuneus | 0.000 | 1.626 |
| 15q11.2del | G_rectus | 3.252 | 4.065 |
| 15q11.2del | G_subcallosal | 1.626 | 3.252 |
| 15q11.2del | G_temp_sup-G_T_transv | 4.878 | 1.626 |
| 15q11.2del | G_temp_sup-Lateral | 2.439 | 1.626 |
| 15q11.2del | G_temp_sup-Plan_polar | 0.813 | 5.691 |
| 15q11.2del | G_temp_sup-Plan_tempo | 3.252 | 1.626 |
| 15q11.2del | G_temporal_inf | 1.626 | 0.813 |
| 15q11.2del | G_temporal_middle | 3.252 | 4.065 |
| 15q11.2del | Lat_Fis-ant-Horizont | 2.439 | 2.439 |
| 15q11.2del | Lat_Fis-ant-Vertical | 2.439 | 1.626 |
| 15q11.2del | Lat_Fis-post | 5.691 | 0.000 |
| 15q11.2del | Pole_occipital | 0.813 | 1.626 |
| 15q11.2del | Pole_temporal | 0.813 | 4.065 |
| 15q11.2del | S_calcarine | 4.065 | 2.439 |
| 15q11.2del | S_central | 2.439 | 2.439 |
| 15q11.2del | S_cingul-Marginalis | 2.439 | 2.439 |
| 15q11.2del | S_circular_insula_ant | 1.626 | 3.252 |
| 15q11.2del | S_circular_insula_inf | 1.626 | 3.252 |
| 15q11.2del | S_circular_insula_sup | 4.065 | 0.813 |
| 15q11.2del | S_collat_transv_ant | 1.626 | 5.691 |
| 15q11.2del | S_collat_transv_post | 2.439 | 3.252 |
| 15q11.2del | S_front_inf | 0.813 | 6.504 |
| 15q11.2del | S_front_middle | 0.813 | 1.626 |
| 15q11.2del | S_front_sup | 0.000 | 0.813 |
| 15q11.2del | S_interm_prim-Jensen | 1.626 | 2.439 |
| 15q11.2del | S_intrapariet_and_P_trans | 1.626 | 1.626 |
| 15q11.2del | S_oc_middle_and_Lunatus | 0.000 | 3.252 |
| 15q11.2del | S_oc_sup_and_transversal | 0.813 | 4.878 |
| 15q11.2del | S_occipital_ant | 0.813 | 0.813 |
| 15q11.2del | S_oc-temp_lat | 3.252 | 1.626 |
| 15q11.2del | S_oc-temp_med_and_Lingual | 2.439 | 3.252 |
| 15q11.2del | S_orbital_lateral | 0.000 | 8.130 |
| 15q11.2del | S_orbital_med-olfact | 3.252 | 4.065 |
| 15q11.2del | S_orbital-H_Shaped | 0.000 | 8.130 |
| 15q11.2del | S_parieto_occipital | 0.813 | 2.439 |
| 15q11.2del | S_pericallosal | 0.813 | 1.626 |
| 15q11.2del | S_postcentral | 1.626 | 3.252 |
| 15q11.2del | S_precentral-inf-part | 0.000 | 0.000 |
| 15q11.2del | S_precentral-sup-part | 2.439 | 1.626 |
| 15q11.2del | S_suborbital | 0.000 | 5.691 |
| 15q11.2del | S_subparietal | 1.626 | 1.626 |
| 15q11.2del | S_temporal_inf | 0.813 | 2.439 |
| 15q11.2del | S_temporal_sup | 0.000 | 6.504 |
| 15q11.2del | S_temporal_transverse | 3.252 | 0.813 |
| 15q11.2dup | G_and_S_frontomargin | 3.311 | 2.649 |
| 15q11.2dup | G_and_S_occipital_inf | 0.662 | 5.298 |
| 15q11.2dup | G_and_S_paracentral | 6.623 | 1.987 |
| 15q11.2dup | G_and_S_subcentral | 5.298 | 1.987 |
| 15q11.2dup | G_and_S_transv_frontopol | 1.987 | 4.636 |
| 15q11.2dup | G_and_S_cingul-Ant | 3.311 | 3.974 |
| 15q11.2dup | G_and_S_cingul-Mid-Ant | 3.311 | 0.662 |
| 15q11.2dup | G_and_S_cingul-Mid-Post | 1.325 | 1.987 |
| 15q11.2dup | G_cingul-Post-dorsal | 1.325 | 2.649 |
| 15q11.2dup | G_cingul-Post-ventral | 2.649 | 3.974 |
| 15q11.2dup | G_cuneus | 1.987 | 1.987 |
| 15q11.2dup | G_front_inf-Opercular | 1.325 | 1.987 |
| 15q11.2dup | G_front_inf-Orbital | 1.325 | 1.987 |
| 15q11.2dup | G_front_inf-Triangul | 2.649 | 3.974 |
| 15q11.2dup | G_front_middle | 3.311 | 3.974 |
| 15q11.2dup | G_front_sup | 0.000 | 3.311 |
| 15q11.2dup | G_Ins_lg_and_S_cent_ins | 3.311 | 2.649 |
| 15q11.2dup | G_insular_short | 2.649 | 1.987 |
| 15q11.2dup | G_occipital_middle | 2.649 | 3.974 |
| 15q11.2dup | G_occipital_sup | 3.974 | 4.636 |
| 15q11.2dup | G_oc-temp_lat-fusifor | 0.662 | 3.311 |
| 15q11.2dup | G_oc-temp_med-Lingual | 0.662 | 2.649 |
| 15q11.2dup | G_oc-temp_med-Parahip | 3.311 | 3.311 |
| 15q11.2dup | G_orbital | 1.325 | 2.649 |
| 15q11.2dup | G_pariet_inf-Angular | 1.325 | 6.623 |
| 15q11.2dup | G_pariet_inf-Supramar | 4.636 | 3.974 |
| 15q11.2dup | G_parietal_sup | 3.974 | 2.649 |
| 15q11.2dup | G_postcentral | 3.311 | 1.987 |
| 15q11.2dup | G_precentral | 2.649 | 2.649 |
| 15q11.2dup | G_precuneus | 3.974 | 1.325 |
| 15q11.2dup | G_rectus | 3.974 | 4.636 |
| 15q11.2dup | G_subcallosal | 0.662 | 6.623 |
| 15q11.2dup | G_temp_sup-G_T_transv | 1.325 | 1.987 |
| 15q11.2dup | G_temp_sup-Lateral | 3.311 | 6.623 |
| 15q11.2dup | G_temp_sup-Plan_polar | 0.662 | 3.974 |
| 15q11.2dup | G_temp_sup-Plan_tempo | 0.662 | 1.325 |
| 15q11.2dup | G_temporal_inf | 3.311 | 4.636 |
| 15q11.2dup | G_temporal_middle | 1.987 | 4.636 |
| 15q11.2dup | Lat_Fis-ant-Horizont | 1.987 | 2.649 |
| 15q11.2dup | Lat_Fis-ant-Vertical | 1.987 | 3.311 |
| 15q11.2dup | Lat_Fis-post | 2.649 | 3.311 |
| 15q11.2dup | Pole_occipital | 4.636 | 3.311 |
| 15q11.2dup | Pole_temporal | 0.662 | 3.311 |
| 15q11.2dup | S_calcarine | 1.987 | 3.311 |
| 15q11.2dup | S_central | 1.325 | 3.311 |
| 15q11.2dup | S_cingul-Marginalis | 0.662 | 1.325 |
| 15q11.2dup | S_circular_insula_ant | 1.987 | 2.649 |
| 15q11.2dup | S_circular_insula_inf | 3.974 | 5.298 |
| 15q11.2dup | S_circular_insula_sup | 2.649 | 3.974 |
| 15q11.2dup | S_collat_transv_ant | 1.325 | 2.649 |
| 15q11.2dup | S_collat_transv_post | 0.662 | 3.974 |
| 15q11.2dup | S_front_inf | 1.987 | 1.987 |
| 15q11.2dup | S_front_middle | 2.649 | 3.311 |
| 15q11.2dup | S_front_sup | 1.325 | 6.623 |
| 15q11.2dup | S_interm_prim-Jensen | 0.662 | 3.311 |
| 15q11.2dup | S_intrapariet_and_P_trans | 2.649 | 4.636 |
| 15q11.2dup | S_oc_middle_and_Lunatus | 1.325 | 3.974 |
| 15q11.2dup | S_oc_sup_and_transversal | 0.662 | 4.636 |
| 15q11.2dup | S_occipital_ant | 1.325 | 3.311 |
| 15q11.2dup | S_oc-temp_lat | 0.662 | 3.311 |
| 15q11.2dup | S_oc-temp_med_and_Lingual | 1.987 | 3.311 |
| 15q11.2dup | S_orbital_lateral | 3.311 | 3.974 |
| 15q11.2dup | S_orbital_med-olfact | 0.000 | 3.311 |
| 15q11.2dup | S_orbital-H_Shaped | 0.000 | 2.649 |
| 15q11.2dup | S_parieto_occipital | 0.000 | 1.325 |
| 15q11.2dup | S_pericallosal | 4.636 | 2.649 |
| 15q11.2dup | S_postcentral | 2.649 | 3.311 |
| 15q11.2dup | S_precentral-inf-part | 1.987 | 1.325 |
| 15q11.2dup | S_precentral-sup-part | 2.649 | 4.636 |
| 15q11.2dup | S_suborbital | 0.662 | 5.960 |
| 15q11.2dup | S_subparietal | 1.987 | 1.987 |
| 15q11.2dup | S_temporal_inf | 0.000 | 2.649 |
| 15q11.2dup | S_temporal_sup | 3.311 | 2.649 |
| 15q11.2dup | S_temporal_transverse | 0.000 | 9.934 |

b) **Left hemisphere for 15q11.2**

| **cnv** | **roi** | **Percentage of positive outliers** | **Percentage of negative outliers** |
| --- | --- | --- | --- |
| 15q11.2del | G_and_S_frontomargin | 1.626 | 1.626 |
| 15q11.2del | G_and_S_occipital_inf | 4.878 | 0.813 |
| 15q11.2del | G_and_S_paracentral | 4.878 | 1.626 |
| 15q11.2del | G_and_S_subcentral | 0.813 | 2.439 |
| 15q11.2del | G_and_S_transv_frontopol | 2.439 | 3.252 |
| 15q11.2del | G_and_S_cingul-Ant | 0.000 | 0.813 |
| 15q11.2del | G_and_S_cingul-Mid-Ant | 2.439 | 2.439 |
| 15q11.2del | G_and_S_cingul-Mid-Post | 1.626 | 2.439 |
| 15q11.2del | G_cingul-Post-dorsal | 2.439 | 1.626 |
| 15q11.2del | G_cingul-Post-ventral | 2.439 | 0.813 |
| 15q11.2del | G_cuneus | 3.252 | 4.065 |
| 15q11.2del | G_front_inf-Opercular | 0.813 | 0.813 |
| 15q11.2del | G_front_inf-Orbital | 4.065 | 4.065 |
| 15q11.2del | G_front_inf-Triangul | 0.813 | 7.317 |
| 15q11.2del | G_front_middle | 0.813 | 1.626 |
| 15q11.2del | G_front_sup | 2.439 | 0.813 |
| 15q11.2del | G_Ins_lg_and_S_cent_ins | 0.813 | 2.439 |
| 15q11.2del | G_insular_short | 2.439 | 2.439 |
| 15q11.2del | G_occipital_middle | 0.813 | 3.252 |
| 15q11.2del | G_occipital_sup | 2.439 | 3.252 |
| 15q11.2del | G_oc-temp_lat-fusifor | 1.626 | 3.252 |
| 15q11.2del | G_oc-temp_med-Lingual | 0.000 | 4.878 |
| 15q11.2del | G_oc-temp_med-Parahip | 4.065 | 2.439 |
| 15q11.2del | G_orbital | 0.000 | 2.439 |
| 15q11.2del | G_pariet_inf-Angular | 0.813 | 4.065 |
| 15q11.2del | G_pariet_inf-Supramar | 2.439 | 1.626 |
| 15q11.2del | G_parietal_sup | 0.813 | 1.626 |
| 15q11.2del | G_postcentral | 2.439 | 1.626 |
| 15q11.2del | G_precentral | 2.439 | 0.813 |
| 15q11.2del | G_precuneus | 1.626 | 0.813 |
| 15q11.2del | G_rectus | 0.813 | 2.439 |
| 15q11.2del | G_subcallosal | 1.626 | 6.504 |
| 15q11.2del | G_temp_sup-G_T_transv | 0.813 | 1.626 |
| 15q11.2del | G_temp_sup-Lateral | 1.626 | 1.626 |
| 15q11.2del | G_temp_sup-Plan_polar | 1.626 | 4.878 |
| 15q11.2del | G_temp_sup-Plan_tempo | 1.626 | 2.439 |
| 15q11.2del | G_temporal_inf | 2.439 | 2.439 |
| 15q11.2del | G_temporal_middle | 2.439 | 1.626 |
| 15q11.2del | Lat_Fis-ant-Horizont | 2.439 | 4.065 |
| 15q11.2del | Lat_Fis-ant-Vertical | 1.626 | 5.691 |
| 15q11.2del | Lat_Fis-post | 4.065 | 3.252 |
| 15q11.2del | Pole_occipital | 4.065 | 4.878 |
| 15q11.2del | Pole_temporal | 0.813 | 4.065 |
| 15q11.2del | S_calcarine | 2.439 | 4.065 |
| 15q11.2del | S_central | 3.252 | 2.439 |
| 15q11.2del | S_cingul-Marginalis | 3.252 | 5.691 |
| 15q11.2del | S_circular_insula_ant | 4.065 | 2.439 |
| 15q11.2del | S_circular_insula_inf | 2.439 | 1.626 |
| 15q11.2del | S_circular_insula_sup | 4.065 | 1.626 |
| 15q11.2del | S_collat_transv_ant | 3.252 | 4.065 |
| 15q11.2del | S_collat_transv_post | 1.626 | 1.626 |
| 15q11.2del | S_front_inf | 2.439 | 4.878 |
| 15q11.2del | S_front_middle | 0.000 | 4.878 |
| 15q11.2del | S_front_sup | 3.252 | 2.439 |
| 15q11.2del | S_interm_prim-Jensen | 2.439 | 3.252 |
| 15q11.2del | S_intrapariet_and_P_trans | 0.813 | 0.813 |
| 15q11.2del | S_oc_middle_and_Lunatus | 0.813 | 8.943 |
| 15q11.2del | S_oc_sup_and_transversal | 0.813 | 3.252 |
| 15q11.2del | S_occipital_ant | 1.626 | 0.000 |
| 15q11.2del | S_oc-temp_lat | 0.813 | 4.065 |
| 15q11.2del | S_oc-temp_med_and_Lingual | 1.626 | 3.252 |
| 15q11.2del | S_orbital_lateral | 0.813 | 7.317 |
| 15q11.2del | S_orbital_med-olfact | 2.439 | 2.439 |
| 15q11.2del | S_orbital-H_Shaped | 0.813 | 2.439 |
| 15q11.2del | S_parieto_occipital | 1.626 | 1.626 |
| 15q11.2del | S_pericallosal | 0.000 | 4.878 |
| 15q11.2del | S_postcentral | 1.626 | 1.626 |
| 15q11.2del | S_precentral-inf-part | 0.813 | 0.813 |
| 15q11.2del | S_precentral-sup-part | 2.439 | 1.626 |
| 15q11.2del | S_suborbital | 2.439 | 1.626 |
| 15q11.2del | S_subparietal | 0.000 | 0.813 |
| 15q11.2del | S_temporal_inf | 0.000 | 5.691 |
| 15q11.2del | S_temporal_sup | 1.626 | 4.878 |
| 15q11.2del | S_temporal_transverse | 1.626 | 0.813 |
| 15q11.2dup | G_and_S_frontomargin | 2.649 | 4.636 |
| 15q11.2dup | G_and_S_occipital_inf | 1.325 | 1.987 |
| 15q11.2dup | G_and_S_paracentral | 5.298 | 1.325 |
| 15q11.2dup | G_and_S_subcentral | 1.987 | 5.298 |
| 15q11.2dup | G_and_S_transv_frontopol | 1.325 | 5.960 |
| 15q11.2dup | G_and_S_cingul-Ant | 1.987 | 1.325 |
| 15q11.2dup | G_and_S_cingul-Mid-Ant | 4.636 | 0.662 |
| 15q11.2dup | G_and_S_cingul-Mid-Post | 3.311 | 1.987 |
| 15q11.2dup | G_cingul-Post-dorsal | 2.649 | 1.987 |
| 15q11.2dup | G_cingul-Post-ventral | 1.987 | 3.311 |
| 15q11.2dup | G_cuneus | 1.325 | 1.987 |
| 15q11.2dup | G_front_inf-Opercular | 3.974 | 3.311 |
| 15q11.2dup | G_front_inf-Orbital | 1.325 | 0.662 |
| 15q11.2dup | G_front_inf-Triangul | 0.662 | 3.311 |
| 15q11.2dup | G_front_middle | 4.636 | 1.325 |
| 15q11.2dup | G_front_sup | 0.662 | 1.987 |
| 15q11.2dup | G_Ins_lg_and_S_cent_ins | 2.649 | 2.649 |
| 15q11.2dup | G_insular_short | 1.325 | 3.974 |
| 15q11.2dup | G_occipital_middle | 0.662 | 2.649 |
| 15q11.2dup | G_occipital_sup | 2.649 | 3.974 |
| 15q11.2dup | G_oc-temp_lat-fusifor | 2.649 | 4.636 |
| 15q11.2dup | G_oc-temp_med-Lingual | 0.000 | 4.636 |
| 15q11.2dup | G_oc-temp_med-Parahip | 1.325 | 1.325 |
| 15q11.2dup | G_orbital | 3.974 | 2.649 |
| 15q11.2dup | G_pariet_inf-Angular | 1.987 | 1.987 |
| 15q11.2dup | G_pariet_inf-Supramar | 2.649 | 3.311 |
| 15q11.2dup | G_parietal_sup | 2.649 | 1.987 |
| 15q11.2dup | G_postcentral | 3.974 | 1.325 |
| 15q11.2dup | G_precentral | 0.662 | 2.649 |
| 15q11.2dup | G_precuneus | 2.649 | 3.311 |
| 15q11.2dup | G_rectus | 3.311 | 3.311 |
| 15q11.2dup | G_subcallosal | 0.000 | 5.298 |
| 15q11.2dup | G_temp_sup-G_T_transv | 1.325 | 1.987 |
| 15q11.2dup | G_temp_sup-Lateral | 1.987 | 3.311 |
| 15q11.2dup | G_temp_sup-Plan_polar | 1.325 | 4.636 |
| 15q11.2dup | G_temp_sup-Plan_tempo | 1.987 | 1.325 |
| 15q11.2dup | G_temporal_inf | 0.662 | 6.623 |
| 15q11.2dup | G_temporal_middle | 0.662 | 3.311 |
| 15q11.2dup | Lat_Fis-ant-Horizont | 0.662 | 3.311 |
| 15q11.2dup | Lat_Fis-ant-Vertical | 0.662 | 3.974 |
| 15q11.2dup | Lat_Fis-post | 3.311 | 4.636 |
| 15q11.2dup | Pole_occipital | 3.974 | 1.987 |
| 15q11.2dup | Pole_temporal | 3.311 | 4.636 |
| 15q11.2dup | S_calcarine | 1.325 | 3.311 |
| 15q11.2dup | S_central | 1.987 | 2.649 |
| 15q11.2dup | S_cingul-Marginalis | 5.298 | 2.649 |
| 15q11.2dup | S_circular_insula_ant | 2.649 | 1.987 |
| 15q11.2dup | S_circular_insula_inf | 3.311 | 2.649 |
| 15q11.2dup | S_circular_insula_sup | 1.325 | 2.649 |
| 15q11.2dup | S_collat_transv_ant | 2.649 | 5.298 |
| 15q11.2dup | S_collat_transv_post | 0.662 | 1.987 |
| 15q11.2dup | S_front_inf | 1.325 | 3.974 |
| 15q11.2dup | S_front_middle | 2.649 | 1.987 |
| 15q11.2dup | S_front_sup | 1.325 | 3.311 |
| 15q11.2dup | S_interm_prim-Jensen | 1.325 | 2.649 |
| 15q11.2dup | S_intrapariet_and_P_trans | 3.311 | 2.649 |
| 15q11.2dup | S_oc_middle_and_Lunatus | 0.000 | 4.636 |
| 15q11.2dup | S_oc_sup_and_transversal | 0.662 | 4.636 |
| 15q11.2dup | S_occipital_ant | 0.662 | 4.636 |
| 15q11.2dup | S_oc-temp_lat | 0.662 | 8.609 |
| 15q11.2dup | S_oc-temp_med_and_Lingual | 3.311 | 1.987 |
| 15q11.2dup | S_orbital_lateral | 1.987 | 4.636 |
| 15q11.2dup | S_orbital_med-olfact | 1.325 | 1.987 |
| 15q11.2dup | S_orbital-H_Shaped | 1.325 | 1.325 |
| 15q11.2dup | S_parieto_occipital | 1.325 | 1.987 |
| 15q11.2dup | S_pericallosal | 1.325 | 3.974 |
| 15q11.2dup | S_postcentral | 4.636 | 5.298 |
| 15q11.2dup | S_precentral-inf-part | 2.649 | 0.662 |
| 15q11.2dup | S_precentral-sup-part | 1.325 | 2.649 |
| 15q11.2dup | S_suborbital | 1.987 | 3.974 |
| 15q11.2dup | S_subparietal | 4.636 | 3.311 |
| 15q11.2dup | S_temporal_inf | 0.662 | 4.636 |
| 15q11.2dup | S_temporal_sup | 1.987 | 4.636 |
| 15q11.2dup | S_temporal_transverse | 0.662 | 5.298 |

**Supplementary Table 10. ANOVA summary of CNVs in Subcortical**

The p-values that remain significant after adjustments for multiple comparisons are highlighted in bold.

| **ROI** | **F-statistic** | **ANOVA P-value** | **Eta Squared** | **Levene Statistic** | **Levene P-value** |
| --- | --- | --- | --- | --- | --- |
| Accumbens | 4.6962 | 0.0091 | 0.0005 | 1.8872 | 0.1515 |
| **Caudate** | **6.9517** | **0.0010** | **0.0007** | **0.6041** | **0.5466** |
| Pallidum | 2.4328 | 0.0878 | 0.0003 | 1.2393 | 0.2896 |
| Putamen | 4.2897 | 0.0137 | 0.0004 | 0.5580 | 0.5724 |
| Thalamus | 1.4135 | 0.2433 | 0.0001 | 0.2161 | 0.8057 |
| Amygdala | 0.8679 | 0.4198 | 0.0001 | 0.2141 | 0.8073 |
| Hippocampus | 2.3630 | 0.0942 | 0.0002 | 0.5844 | 0.5574 |
| **Thickness** | **10.6211** | **0.0000** | **0.0011** | **0.4963** | **0.6088** |
| Intracranial-Vol | 4.3819 | 0.0125 | 0.0005 | 0.0275 | 0.9729 |
| **SurfaceArea** | **7.9769** | **0.0003** | **0.0008** | **0.2323** | **0.7927** |

**Supplementary Table 11. Associations of CNV with Global and Subcortical Brain Morphology**

The p-values that remain significant after adjustments for multiple comparisons are highlighted in bold

| **cnv** | **roi** | **t-stat** | **p-value** | **cohens d** | **ci_lower** | **ci_upper** | **median_hc** | **median_c** |
| --- | --- | --- | --- | --- | --- | --- | --- | --- |
| 15q11.2dup | Accumbens | -0.4506 | 0.6530 | -0.0390 | -0.2007 | 0.1227 | -0.0812 | -0.1703 |
| **15q11.2dup** | **Caudate** | **-2.8852** | **0.0045** | **-0.2408** | **-0.4026** | **-0.0791** | **-0.1124** | **-0.3397** |
| 15q11.2dup | Pallidum | -1.1811 | 0.2394 | -0.0991 | -0.2608 | 0.0627 | -0.0632 | -0.1730 |
| 15q11.2dup | Putamen | -1.5647 | 0.1198 | -0.1307 | -0.2925 | 0.0310 | -0.0740 | -0.2669 |
| 15q11.2dup | Thalamus | -1.3064 | 0.1934 | -0.1066 | -0.2683 | 0.0552 | -0.0965 | -0.2394 |
| 15q11.2dup | Amygdala | -1.0512 | 0.2949 | -0.0920 | -0.2538 | 0.0697 | -0.0955 | -0.2898 |
| 15q11.2dup | Hippocampus | -1.8377 | 0.0681 | -0.1446 | -0.3063 | 0.0171 | -0.0532 | -0.1689 |
| 15q11.2dup | Thickness | -2.8250 | 0.0054 | -0.2191 | -0.3809 | -0.0574 | 0.0150 | -0.1485 |
| **15q11.2dup** | **Intracranial-Vol** | **-2.9779** | **0.0034** | **-0.2430** | **-0.4047** | **-0.0812** | **-0.0876** | **-0.4472** |
| **15q11.2dup** | **SurfaceArea** | **-3.2066** | **0.0016** | **-0.2876** | **-0.4493** | **-0.1258** | **-0.0477** | **-0.8341** |
| **15q11.2del** | **Accumbens** | **-3.4247** | **0.0008** | **-0.2777** | **-0.4572** | **-0.0982** | **-0.0812** | **-0.3212** |
| 15q11.2del | Caudate | -2.6938 | 0.0081 | -0.2141 | -0.3936 | -0.0346 | -0.1124 | -0.3217 |
| 15q11.2del | Pallidum | -2.0675 | 0.0408 | -0.1702 | -0.3497 | 0.0093 | -0.0632 | -0.2190 |
| 15q11.2del | Putamen | -2.7178 | 0.0075 | -0.2266 | -0.4061 | -0.0471 | -0.0740 | -0.3326 |
| 15q11.2del | Thalamus | -1.0585 | 0.2919 | -0.0994 | -0.2789 | 0.0801 | -0.0965 | -0.2720 |
| 15q11.2del | Amygdala | -0.6997 | 0.4854 | -0.0650 | -0.2445 | 0.1145 | -0.0955 | -0.2005 |
| 15q11.2del | Hippocampus | -1.2272 | 0.2221 | -0.1190 | -0.2985 | 0.0605 | -0.0532 | -0.1319 |
| **15q11.2del** | **Thickness** | **3.9503** | **0.0001** | **0.3431** | **0.1635** | **0.5226** | **0.0150** | **0.3912** |
| 15q11.2del | Intracranial-Vol | 0.2904 | 0.7720 | 0.0260 | -0.1534 | 0.2055 | -0.0876 | -0.0559 |
| 15q11.2del | SurfaceArea | -2.0089 | 0.0468 | -0.1810 | -0.3605 | -0.0015 | -0.0477 | -0.4120 |
